# Romosozumab Safely Restores Bone Mass in Multiple Myeloma via Osteoblast Reprogramming: A Phase IIa Study

**DOI:** 10.64898/2026.07.02.26357196

**Authors:** Betty Gration, Ryan C. Chai, Shelley G. Young, C. Marcelo Sergio, Elena Skorokhodova, James T. Smith, Ariel Castro-Martinez, Alessandra Bray, Xufeng Lin, Calysta Yan, Joanna Kao, Jacinta Perram, Sam Lai, Louis Lau, Katherine N. Weilbaecher, John Moore, Nicholas Pocock, Christine L. Chaffer, Jackie Center, Tri Giang Phan, Georgia McCaughan, Peter I. Croucher

## Abstract

Multiple myeloma causes devastating osteolytic bone disease. Current antiresorptive therapies slow bone loss but fail to rebuild the skeleton. Consequently, patients continue to fracture and suffer the associated morbidity and mortality. Targeting the Wnt inhibitor sclerostin, with romosozumab, increases bone mass in osteoporosis but has not been leveraged in cancer. We hypothesised that romosozumab would safely restore bone mass in multiple myeloma. In a murine model of myeloma, romosozumab was safe, demonstrating no impact on tumour progression while significantly increasing bone density. We subsequently conducted a Phase IIa proof-of-concept study in 12 multiple myeloma patients refractory to bisphosphonate therapy. Romosozumab was safe, it was well-tolerated and did not promote clinical or clonal myeloma progression. Treatment induced an early, significant rise in serum bone formation markers whilst resorption remained unchanged. This was coupled with significant gains in bone mineral density throughout the skeleton. Additionally, we observed radiological evidence of repair to pre-existing osteolytic lesions and, critically, a reduction in the skeletal morbidity rate from 2.5 to 0.11 events per patient-year. Longitudinal single-cell transcriptomics revealed that romosozumab transiently reprograms the osteoblast lineage to upregulate matrix synthesis and mineralisation programmes. These findings demonstrate that sclerostin inhibition safely rebuilds bone, providing clinical and mechanistic rationale for further randomised studies to restore bone health in myeloma patients.

## INTRODUCTION

Multiple myeloma is a plasma cell malignancy that causes devastating bone disease characterised by osteolytic lesions, bone loss, pain, and fractures. Osteolytic bone disease is present at diagnosis in 80% of patients^1,2^. Fractures lead to substantial morbidity and are associated with a twofold increased risk of death^3^. The bone disease results from uncoupling of normal bone remodelling, with osteoclast-mediated resorption markedly increased while osteoblast-mediated bone formation is simultaneously suppressed. Current standard-of-care antiresorptive therapies, zoledronic acid or the RANK ligand antibody denosumab^4,5^, slow further bone loss but do not restore lost bone, rebuild bone strength, or repair existing osteolytic lesions. Consequently, 45% of patients sustain skeletal-related events despite antiresorptive therapy^6^. There is a critical need for bone anabolic strategies capable of actively rebuilding the skeleton in patients with myeloma.

The molecular basis of osteoblast suppression is well characterised. Myeloma cells and cells in the local microenvironment produce inhibitors of bone formation including the Wnt antagonists dickkopf-1, secreted frizzled-related proteins 2 and 3, activin A, and interleukins 3 and 7^7–11^. Epigenetic reprogramming of mesenchymal stromal cells also drives sustained, long-term osteoblast suppression^12,13^. Targeting these pathways arrests myeloma bone disease in experimental models, yet none has translated successfully into clinical practice^14–19^. We and others discovered that sclerostin, an osteocyte-derived inhibitor of canonical Wnt signalling, is elevated in myeloma patient serum, correlates with markers of bone destruction, and when neutralised in preclinical models increases bone formation, bone mass, and critically, bone strength^20–24^. Sclerostin is generally not produced by myeloma cells but originates exclusively from osteocytes embedded within bone matrix^22^. This local tissue-restricted biology circumvents the longstanding concern, and principal barrier to translation, that broad Wnt pathway activation could drive tumour progression. Moreover, because sclerostin is a product of the native bone microenvironment rather than a tumour-specific factor, anti-sclerostin therapy would be expected to benefit all patients with myeloma bone disease regardless of molecular subtype.

Romosozumab is a humanised monoclonal antibody that neutralises sclerostin and is licensed for osteoporosis^25–27^. It uniquely stimulates new bone formation and suppresses bone resorption, producing rapid, substantial gains in bone mass^26,27^. Unlike antiresorptive agents, romosozumab has a lower risk of osteonecrosis of the jaw and is not associated with the significant rebound bone loss sometimes seen with denosumab discontinuation^28–32^. Romozosumab is not licensed for any cancer-related bone disease, yet these characteristics make it a compelling candidate.

We hypothesised that romosozumab would be safe in patients with multiple myeloma, promote new bone formation, repair established osteolytic lesions and prevent new skeletal-related events (SRE). We first demonstrated that romosozumab had no effect on myeloma burden, time to morbidity of other safety concerns, yet was still able to stimulate bone formation and increase bone mass in a murine model of myeloma. This provided the rationale to conduct a phase IIa proof-of-concept clinical study of romosozumab in patients with multiple myeloma who had a new SRE despite treatment with zoledronic acid. Incorporation of longitudinal single-cell RNA sequencing of bone and myeloma cells was used to define the underlying molecular mechanisms. Romosozumab was safe, well tolerated, and critically did not accelerate myeloma progression. Treated patients demonstrated increased biochemical markers of bone formation, significant gains in bone mineral density, radiological repair of pre-existing lytic lesions and a profound reduction in skeletal morbidity rate.

These findings represent a fundamental shift in the approach to treating myeloma bone disease, providing the mechanistic rationale, clinical proof of concept and safety data to support romosozumab evaluation in randomised controlled trials, with the potential to transform skeletal outcomes for patients with myeloma.

## RESULTS

### Romosozumab is Safe and Increases Bone Mineral Density in a Murine Model of Myeloma

To determine whether romosozumab affects myeloma burden and disease progression, and whether it could prevent myeloma bone disease, we treated naïve mice, or mice bearing syngeneic 5TGM1 murine myeloma cells, with romosozumab weekly for 3 weeks (Fig. 1a). Flow cytometric analysis showed that romosozumab did not change 5TGM1 myeloma burden in bone marrow or spleen (Fig. 1b). Romosozumab treatment also had no effect on the time to ethical endpoint (Fig. 1c). Whilst control-treated 5TGM1 mice exhibited progressive weight loss and lower final body weight, romosozumab treatment reduced myeloma-induced weight loss. (Suppl. Fig. 1a). Romosozumab treatment also reduced experimental paralysis degree (EPD) scores, a measure of animal well-being (Suppl. Fig. 1b).

**Figure 1:**
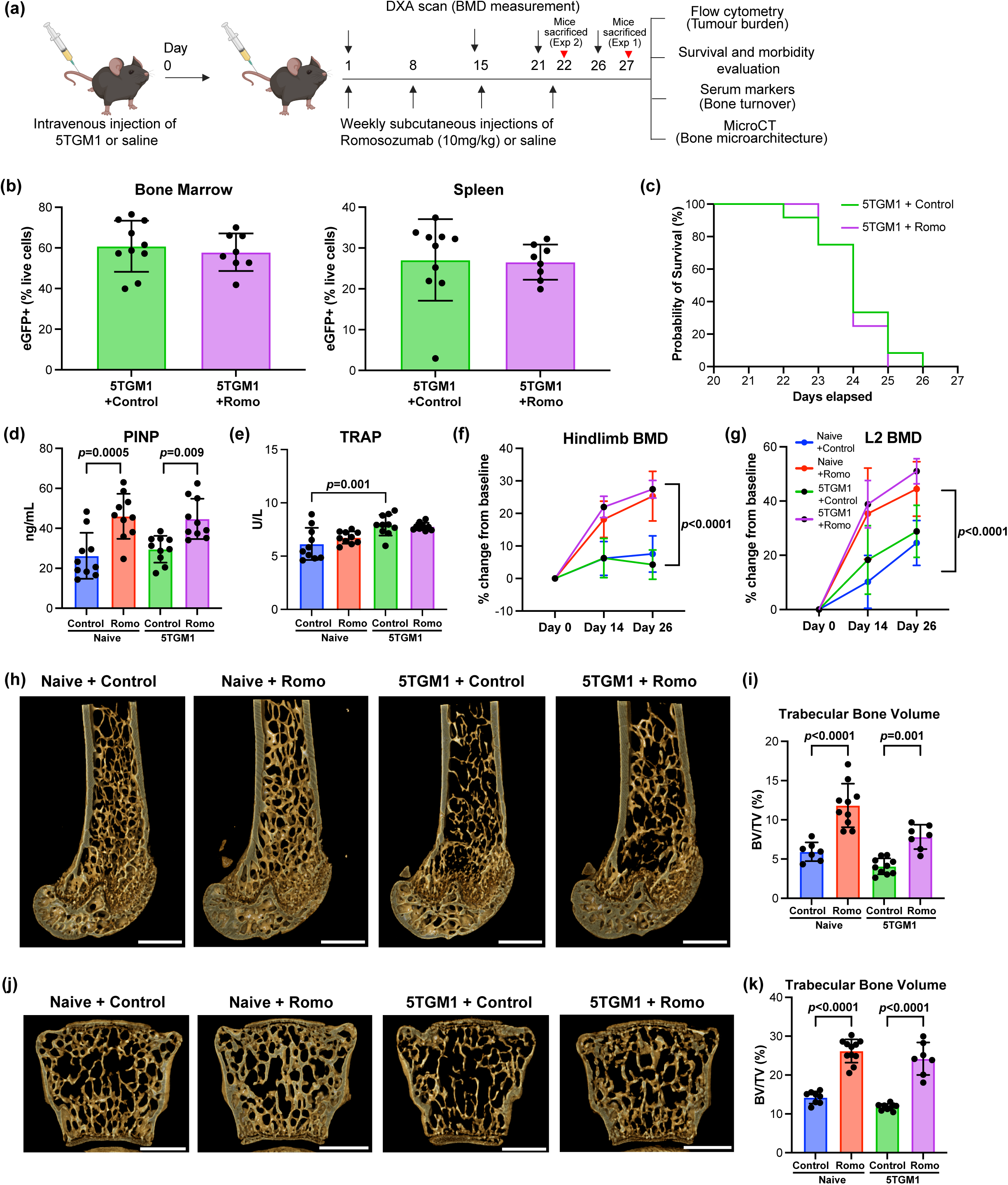
Romosozumab does not increase myeloma burden but increases bone density in murine model of myeloma **(a)** Experimental schematic of the 5TGM1 multiple myeloma murine model and romosozumab treatment timeline. **(b)** Quantification of tumour burden determined by the percentage of live 5TGM1 myeloma cells, within the bone marrow and spleen. **(c)** Kaplan-Meier survival curves of tumour-bearing mice receiving either weekly romosozumab or vehicle control. Quantification of serum bone turnover markers representing **(d)** bone formation (PINP) and **(e)** bone resorption (TRAP) activity. Longitudinal dual-energy X-ray absorptiometry (DXA)-derived bone mineral density (BMD) measurements in the **(f)** hindlimb and **(g)** L2 vertebra, expressed as percentage change from baseline. **(h)** Representative 3D micro-computed tomography (microCT) reconstructions of distal femoral trabecular bone across treatment groups; scale bars = 1 mm. **(i)** MicroCT quantification of distal femoral trabecular bone volume fraction (BV/TV). **(j)** Representative 3D microCT reconstructions of L2 vertebrae across treatment groups; scale bars = 1 mm. **(k)** MicroCT quantification of L2 vertebral trabecular BV/TV. Data are presented as mean ± SD. Statistical significance was evaluated using a two-way ANOVA with Šídák’s multiple comparisons test for **(f, g),** unpaired Welch’s *t*-test for **(b)** and a one-way ANOVA with Tukey’s multiple comparisons test for all other parameters.

Romosozumab treatment increased the serum bone formation marker procollagen type 1 N-terminal propeptide (PINP) in both naïve and myeloma-bearing mice (Fig. 1d). Serum tartrate-resistant acid phosphatase (TRAP), a marker of osteoclastic bone resorption, was increased in myeloma bearing mice compared to control (Fig. 1e). Romosozumab treatment had no effect on serum TRAP in either naïve or tumour bearing 5TGM1-bearing mice. The romosozumab-induced increase in bone formation was associated with an increase in bone mineral density (BMD) in both the hindlimb and L2 vertebrae in naïve and myeloma-bearing mice (Fig. 1f,g). Micro-computed tomography (microCT) analysis showed that romosozumab increased trabecular bone volume, thickness and number in both the femur and vertebrae in naïve and myeloma-bearing mice (Fig. 1h-k; Suppl. Fig. 1c,d). Trabecular separation was increased in myeloma-bearing mice in the femur but not vertebra, whereas romosozumab decreased separation in the vertebra of both naïve and myeloma-bearing mice (Suppl. Fig. 1c,d). Analysis of cortical bone in the femur showed that treatment increased both cortical volume and thickness (Suppl. Fig. 1e). The effects on bone density and femoral bone microarchitecture were replicated in independent experiments (Suppl. Table 1).

Together, these preclinical studies demonstrate that romosozumab does not increase myeloma burden or disease progression yet increases bone formation and stops myeloma-induced bone disease.

### A Phase IIa, Proof-of-Concept Study of Romosozumab in Myeloma Patients

Based on the demonstration that romosozumab does not increase myeloma burden in mice we initiated an open-label phase IIa proof-of-concept study of romosozumab in patients with multiple myeloma that had experienced an SRE despite zoledronic acid (ACTRN12624000027516).

#### Patient Characteristics

Twelve patients with multiple myeloma were enrolled in the study (Table 1). The median age was 69 years (IQR: 54, 74.25) and four patients (33%) were male. All patients were heavily pre-treated with bisphosphonates; seven patients (58%) had received more than three years of bisphosphonate therapy prior to enrolment.

**Table 1:** Patient Characteristics at Baseline.

| Characteristic |  |
| --- | --- |
| Median age (years) (IQR) | 69.0 (54.0, 74.3) |
| BMI, median (IQR) | 26.5 (23.7, 31.7) |
| Gender Male, n (%) | 4 (33%) |
| ISS, median (IQR) | 1.5 (1.0, 2.0) |
| Current line of therapy, median (IQR) | 1 (1.0, 2.5) |
| Duration of bisphosphonate use, n (%) |  |
| <1 year | 2 (17%) |
| 1-3 years | 3 (25%) |
| >3 years | 7 (58%) |
| Median Bone Mineral Density T Score (IQR) |  |
| Lumbar spine | 0.3 (-0.9, 0.9) |
| Total hip | -1.5 (-2.1, -0.7) |
| Number of Osteolytic lesions, n (%) |  |
| 0 | 1 (8%) |
| 1-2 | 1 (8%) |
| 3-5 | 1 (8%) |
| 5-10 | 0 (0%) |
| 11-20 | 3 (25%) |
| 21-50 | 3 (25%) |
| >50 | 3 (25%) |
| Median serum PINP (ug/L) (IQR) | 34 (26.3, 58.3) |
| Median serum beta-CTX (ug/L) (IQR) | 240 (165.8, 402.5) |

At baseline, the median BMD T-score was 0.3 (IQR: –0.93, 0.9) at the lumbar spine and –1.5 (IQR: –2.1, –0.7) at the total hip. Consistent with heavy prior antiresorptive use, baseline median serum concentrations of bone turnover markers were low; procollagen type 1 N-terminal propeptide (PINP) was 34ug/L (IQR: 26.3, 58.3) and beta-C terminal telopeptide (beta-CTX) was 240ug/L (IQR: 165.8, 402.5).

Patients demonstrated a significant burden of active and historical myeloma bone disease on baseline imaging. All 12 patients (100%) had a history of at least one SRE (defined as new fractures, spinal cord compression or need for radiotherapy or surgery)^1^, prior to enrolment with a total of 98 historical SREs in the cohort. The median number of prior SREs per patient was 5.5 (IQR: 1.75, 15.5) with five patients (41.7%) having had more than 10 SREs prior to initiating romosozumab. Nine patients (75%) presented with more than 10 radiologically identifiable osteolytic lesions at baseline, including three patients (25%) with greater than 50 lesions. The median International Staging System (ISS) score was 1.5 (IQR: 1, 2.5). Patients had received a median of one prior lines of therapy (IQR: 1, 2.5). Myeloma treatment regimens differed but no patient changed systemic therapy whilst on study.

#### Safety and Adverse Events

Romosozumab was well-tolerated with a safety profile consistent with the underlying disease and prior treatment histories (Table 2). The only treatment-emergent adverse events (TEAEs) considered related to the study drug were grade 1-2 injection site reactions (mild-moderate erythema).

**Table 2:** Adverse Events.

| **Adverse event** | **Grade 1-2,**<br>**n (%)** | **Grade ≥3,**<br>**n (%)** | **Treatment related**<br>**n (%)** |
| --- | --- | --- | --- |
| Infection | 5 (42%) | 1 (8%) | 0 (0%) |
| Arthralgia | 1 (8%) | 0 (0%) | 0 (0%) |
| Increased perspiration | 1 (8%) | 0 (0%) | 0 (0%) |
| Rash | 2 (17%) | 0 (0%) | 0 (0%) |
| Seizure | 0 (0%) | 1 (8%) | 0 (0%) |
| Injection site reaction | 4 (33%) | 0 (0%) | 4 (33%) |
| Nausea and vomiting | 1 (8%) | 0 (0%) | 0 (0%) |
| Diarrhoea | 1 (8%) | 0 (0%) | 0 (0%) |
| Abdominal pain | 1 (8%) | 0 (0%) | 0 (0%) |
| Bone pain | 5 (42%) | 1 (8%) | 0 (0%) |
| Cramp | 1 (8%) | 1 (8%) | 0 (0%) |
| Skeletal-related event | 1 (8%) | 0 (%) | 0 (0%) |
| Neutropenia | 1 (8%) | 1 (8%) | 0 (0%) |
| Discontinued study | 0 (0%) | 2 (17%) | 0 (0%) |
| Progressive<br>myeloma | 1 (8%) | 1 (8%) | 0 (0%) |
| Myeloid neoplasm | 1 (8%) | 1 (8%) | 0 (0%) |

One patient experienced a grade 3 seizure. Further evaluation deemed this was secondary to a historic infarct and unrelated to romosozumab. Other non-serious adverse events seen were typical of a heavily pre-treated myeloma cohort, including cytopenias, bone pain and respiratory tract infections. These did not require dose modification or trial discontinuation.

Serious adverse events (SAEs) were rare and not attributable to romosozumab therapy. Two patients discontinued the trial due to Grade 5 (fatal) events unrelated to romosozumab. One was secondary to refractory multiple myeloma (enrolled on study at progression and failed to respond to further anti-neoplastic therapy) and the second a therapy-related myeloid neoplasm, presumed related to prior lenalidomide.

There were no reports of adverse events of special interest previously monitored in large phase III romosozumab osteoporosis trials^29–32^. No patients developed hypocalcemia, hyperostosis, osteoarthritis, osteonecrosis of the jaw, atypical fracture or cardiovascular events.

Patients were monitored for disease progression via serum biomarkers and feedback from treating physicians. All patients remained within the same, or improved, response criteria throughout the 12-month treatment period as per criteria set out by the International Myeloma Working Group (IMWG)^33^ (Figure 4e; Suppl. Table 2). At enrollment, two patients had progressive disease and continued to progress on study. One, as discussed above, failed to respond to further anti-neoplastic therapy. The second, had an asymptomatic lesion identified on an off-protocol PET-CT that had not been visualised on screening CT skeletal survey acknowledging the imaging modalities are not directly comparable. This single asymptomatic lesion was managed with radiotherapy and there was no change in systemic treatment. However, as per definition, this was recorded as progression and an SRE.

### Romosozumab Increases Bone Formation and Bone Mineral Density

#### Bone Turnover Markers

Romosozumab significantly increased serum levels of PINP, a marker of new bone formation (overall ANOVA F = 5.52, p < 0.001) (Fig. 2b). After 1 month (a single dose) of treatment, the estimated marginal mean (EMM) PINP rose to 88.7% from baseline (95% CI: 25.3%, 152.0%, p = 0.003). This increase in EMM peaked at 2 months, 94.9% above baseline (95% CI: 27.4%, 162.4%, p = 0.003). From 3 months the PINP began to decrease; at 3 months it was no longer significantly different from baseline after strict multiple testing corrections, and by 6 months had returned to near-baseline levels (EMM –3.7%, p = 0.998).

**Figure 2:**
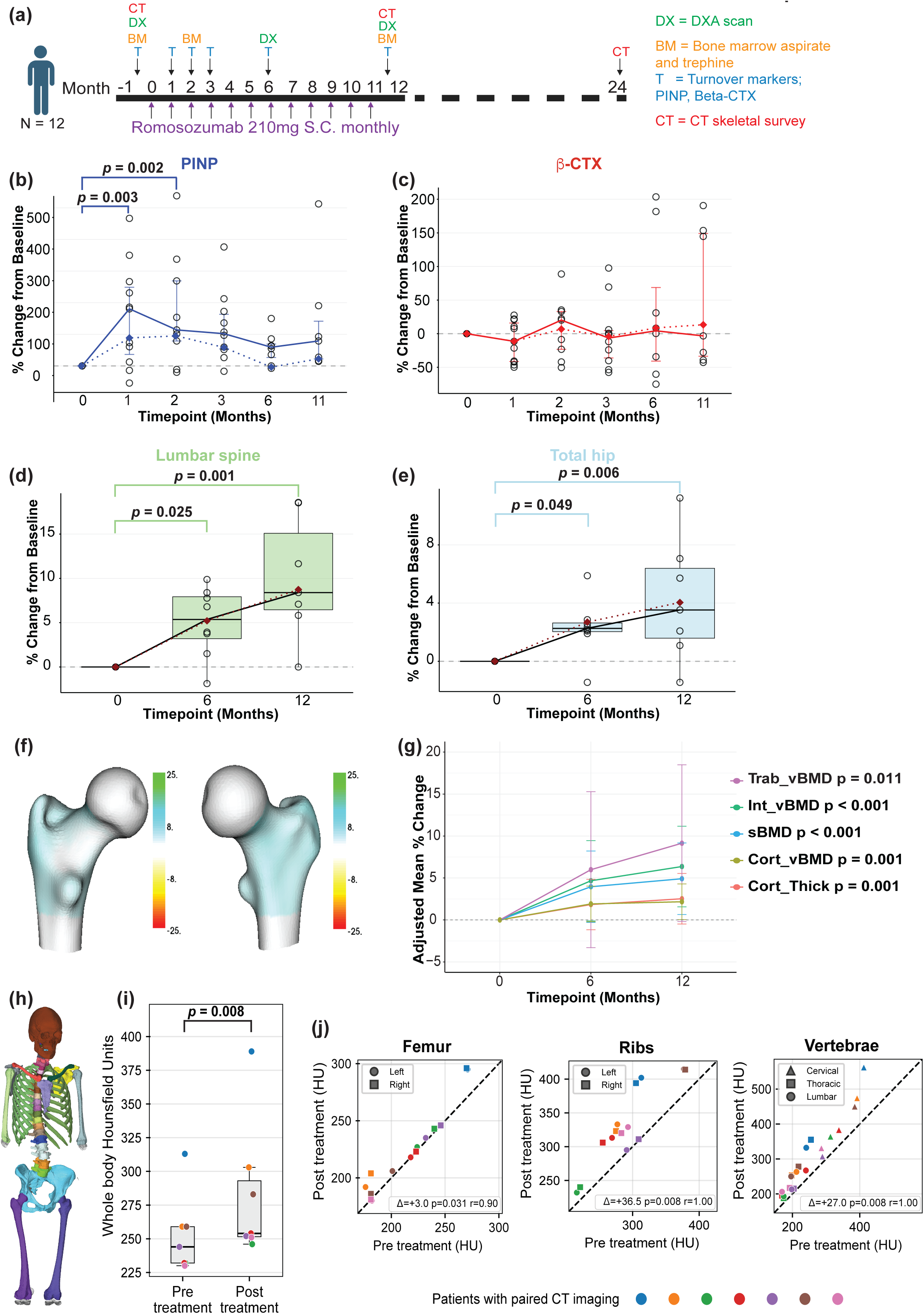
Romosozumab increases bone formation and bone mineral density in MM patients **(a)** Trial schematic and timing of procedures and sampling. **(b)** Percentage change in PINP from baseline. Solid line represents median values, circles are individual patient values and dotted lines are estimated marginal means derived from the linear mixed-effects model. **(c)** Percentage change in beta-CTX from baseline. Lines and circles are described in b. **(d)** Percentage change in aBMD determined by 2D-DXA at the lumbar spine. Lines and circles are described in b. Box and whiskers represent median and range **(e)** Percentage change in aBMD determined by 2D-DXA at the total hip. Annotation as in d. **(f)** Representative images of a pair of femurs showing the median percentage change in the 3D shaper measure of sBMD (surrogate of bone strength and fracture resistance) from baseline to 12-months. Scale bars show differences in sBMD with an increase shown in blue-green and a decrease in yellow-red. **(g)** Adjusted mean percentage change in cortical and trabecular compartments from baseline as calculated using 3D shaper. p-values are for the overall effect (ANOVA). Mean and range are shown at each timepoint. **(h)** Skellytour bone segmentation from a representative study patient. **(i)** Box and whisker plot of median whole-body bone radiodensity (measured in HU) pre and post treatment. Each coloured spot denotes the median value from an individual patient. **(j)** Scatter plot comparing regional whole-bone radiodensity (measured in HU) pre– and post-treatment. The dashed diagonal line represents the line of identity; patient median values (represented as coloured shapes) falling above this line demonstrate an increase in radiodensity post-treatment. Specific anatomical sub-regions or laterality is represented by a different shape.

By contrast, the bone resorption marker beta-CTX did not demonstrate any significant overall change across the 12-month treatment period (F = 0.71, p = 0.620) (Fig. 2c). This indicates that new bone formation was uncoupled from bone resorption without a significant increase in overall suppression of resorptive activity.

#### Areal Bone Mineral Density

The increase in bone formation was associated with significant increases in areal bone mineral density (aBMD) at all evaluated sites both at 6 and 12 months. Linear mixed-effects modelling revealed a significant overall increase in aBMD with time on treatment at both the lumbar spine (F = 10.87, p = 0.002) and total hip (F = 6.96, p = 0.009) (Figs 2d, e). At the lumbar spine, EMM aBMD increased from a baseline of 1.198 g/cm2 to 1.261 g/cm2 at 6 months, representing an increase of 5.2% (95% CI: 1.3%, 9.2%, p = 0.025). This trajectory continued to the end of the treatment period with an EMM of 1.303 g/cm2 at 12-months representing an increase of 8.7% from baseline (95% CI: 4.6%, 12.9%, p < 0.001). This significant improvement was also noted at the total hip where EMM aBMD increased from a baseline of 0.888 g/cm2 to 0.912 g/cm2 at 6 months (2.7% increase, 95% CI: 0.4%, 5.0%, p = 0.049) and 0.924 g/cm2 by 12 months (4.0% increase, 95% CI: 1.6%, 6.5%, p = 0.007).

#### Volumetric and Compartmental Microarchitecture

Given the changes seen in both trabecular and cortical bone compartments in mice treated with romosozumab, we performed a 3-dimensional compartmental analysis of BMD data using 3D-SHAPER^34,35^. This exploratory analysis demonstrated robust and statistically significant improvements across all bone compartments over time (Fig. 2g). Integral volumetric BMD (Int_vBMD) showed a clear overall increase (F = 14.94, p < 0.001), rising by 4.7% (95% CI: 1.9%, 7.4%, p < 0.001) after 6 months and 6.4% (95% CI: 3.4%, 9.3%, p < 0.001) by 12 months. Trabecular vBMD (Trab_vBMD) exhibited the largest relative increase from a baseline EMM of 125.1 mg/cm3 to 136.6 mg/cm3 after 12 months (F = 5.2, p = 0.011). The rise of 6% at 6 months was not significant after adjustment but there was a statistically significant rise of 9.1% at 12 months (95% CI: 2.3%, 16.0%, p = 0.008). In the cortical compartment both cortical thickness and density were increased. Cortical vBMD (Cort_vBMD) increased by 2.2% (95% CI: 0.85, 3.6%; p = 0.002) at 12 months and cortical thickness increased from EMM of 1.86 mm to 1.91 mm; a 2.5% increase (95% CI: 1.5%, 2.7%, p = 0.002). Furthermore, we saw highly significant improvements in the cortical surface BMD (sBMD) over time (F = 20.4, p < 0.001) as shown by the 3D model of the proximal femur (Fig 2g). This parameter is the product of cortical thickness and volumetric density and therefore acts as a surrogate for cortical bone strength and reduced resistance to fracture^35,36^. This improved resistance was seen at both the 6-month (4% increase, 95% CI: 2.1%, 5.8%, p<0.001) and 12-month (4.9% increase, 95% CI: 2.9%, 6.9%, p < 0.001) assessments relative to baseline.

#### Radiodensity analysis of the skeleton beyond the hip and spine

Baseline CT imaging showed that skeletal disease was not restricted to the hip and spine within our cohort. We therefore performed further exploratory analyses to examine the ability of romosozumab to increase bone mass at other skeletal sites. To do this we utilised Skellytour, an AI-assisted segmentation tool^37,38^, to extract a measure of radiodensity (Hounsfield Units, HU) from all bones proximal of the knee and elbow (as shown by representative patient segmentation in Fig. 2h) on paired pre-and post-treatment CT scans. Across the evaluable cohort (n = 7), the median whole body radiodensity increased significantly following 12 months of romosozumab with a median increase of 22.0 HU (IQR: 17.5, 34.0, p = 0.008) (Fig 2i).

When assessing specific whole-bone regions (Fig 2j), significant increases in median HU were observed at the bilateral femora (increase 3.0HU, IQR: 0.8, 12.3, p = 0.031), bilateral ribs (increase 36.5HU, IQR: 27.0, 47.8, p = 0.008) and all vertebrae (increase 27.0HU, IQR: 22.0, 55.5, p = 0.008). These data suggest that romosozumab is able to increase bone density in patients with myeloma across multiple bone sites.

### Radiological Lesion Repair

To determine whether romosozumab could aid in the repair of pre-existing lesions, we performed retrospective radiological review of paired pre– and post-treatment CT skeletal surveys. This demonstrated measurable structural improvements within a subset of patients. Seven measurable lesions were identified and assessed in detail. Four of the seven lesions (57%) demonstrated a reduction in cross-sectional size following 12 months of romosozumab treatment (Fig. 3a, b). Lesions shown to undergo repair had a rim of new sclerotic bone on the post-treatment scans that was further assessed using high-resolution spatial mapping of radiodensity (HU) (Fig. 3c). This revealed new bone formation, and increase in radiodensity, to be primarily occurring at the lesion periphery on existing bone surfaces, with minimal radiodensity changes observed within the central region of the lesion (Fig. 3d). This is consistent with new bone being deposited on existing bone surfaces rather than via de novo bone formation.

**Figure 3:**
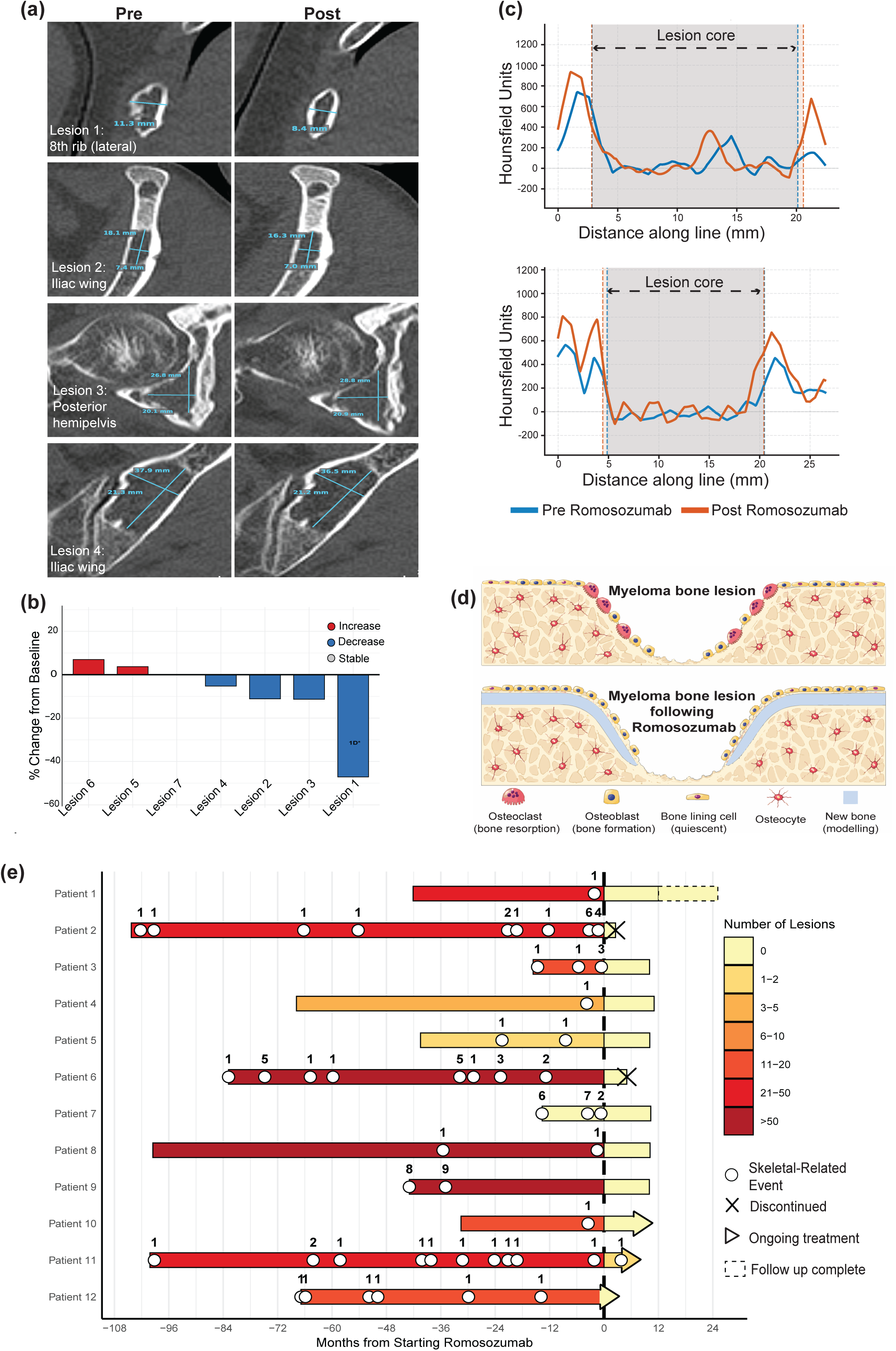
Radiological Lesion Repair and Prevention of Skeletal-Related Events. **(a)** CT images of lesions pre-(left) and post-(right) romosozumab treatment; (i) is 8th rib, (ii) left iliac wing, (iii) posterior right hemipelvis and (iv) left iliac wing. **(b)** Waterfall plot demonstrating percentage change in lesion size after 12 months of romosozumab treatment relative to baseline. The lesion with measurement in only 1 dimension is denoted. **(c)** Radiodensity across representative lesions pre– and post-romosozumab treatment. **(d)** Schematic illustrating the proposed mechanism of lesion repair with romosozumab treatment. **(e)** Swimmer plot representing the burden of skeletal disease across the patient cohort before (left of dashed line) and after starting romosozumab treatment (right of dashed line). The number of lytic lesions are shown in the heatmap. The timing and number of new skeletal related events are identified as circles on the individual patient timelines.

### Romosozumab Prevents Skeletal-Related Events

Our patient cohort had a significant burden of skeletal disease at baseline as evidenced by the cumulative history of 98 SREs prior to study commencement (Fig. 3e). Looking at the 12 months immediately prior versus the 12 months on romosozumab; the median number of SREs pre-treatment was 1.0 per patient equating to a skeletal morbidity rate (SMR) of 2.5 events per patient-year. This is a dramatic contrast to the post-treatment median of 0 SREs with an exposure-adjusted SMR of just 0.11 events per patient-year. The single SRE reported on study was identified during intensive, off-protocol PET-CT monitoring and led to prophylactic radiotherapy administered to an asymptomatic lesion. This suggests romosozumab is stabilizing skeletal disease in this patient cohort.

### Romosozumab Treatment Does Not Accelerate Myeloma Progression

To determine whether romosozumab promotes myeloma progression we performed single-cell B-cell receptor (BCR) and RNA sequencing of cells isolated from paired bone trephines and marrow aspirates taken before, 2 months and 11 months after starting romosozumab. Unsupervised clustering identified distinct myeloid, lymphoid and non-hematopoietic lineages, consistent with the landscape of cells recently described in the endosteal compartment of normal human bone^39^ (Fig. 4a).

Mapping of both light– and heavy-chains enabled identification of both the dominant myeloma clone (clonotype 1), minor clones (clonotypes 2-5) and individual cells expressing unique BCRs, which are normal plasma cells (denoted as other clonotypes) (Fig. 4b). Longitudinal cell composition analysis in the bone trephines showed that the frequency of dominant clones remained stable at 2 months but decreased at 11 months. In the bone marrow aspirate the dominant myeloma clone was decreased at both 2-and 11-month timepoints (Fig. 4c; Suppl. Fig. 3a,b). A small number of minor clones were seen at each time point in both trephine and aspirate biopsies although these represent <5% of all plasma cells. Reductions in the dominant clone frequency at 2 and 11 months were associated with increases in the proportion of normal plasma cells (Fig. 4b,c). The decrease in dominant myeloma clones likely reflects the therapeutic efficacy of concurrent, ongoing anti-myeloma treatment.

**Figure 4:**
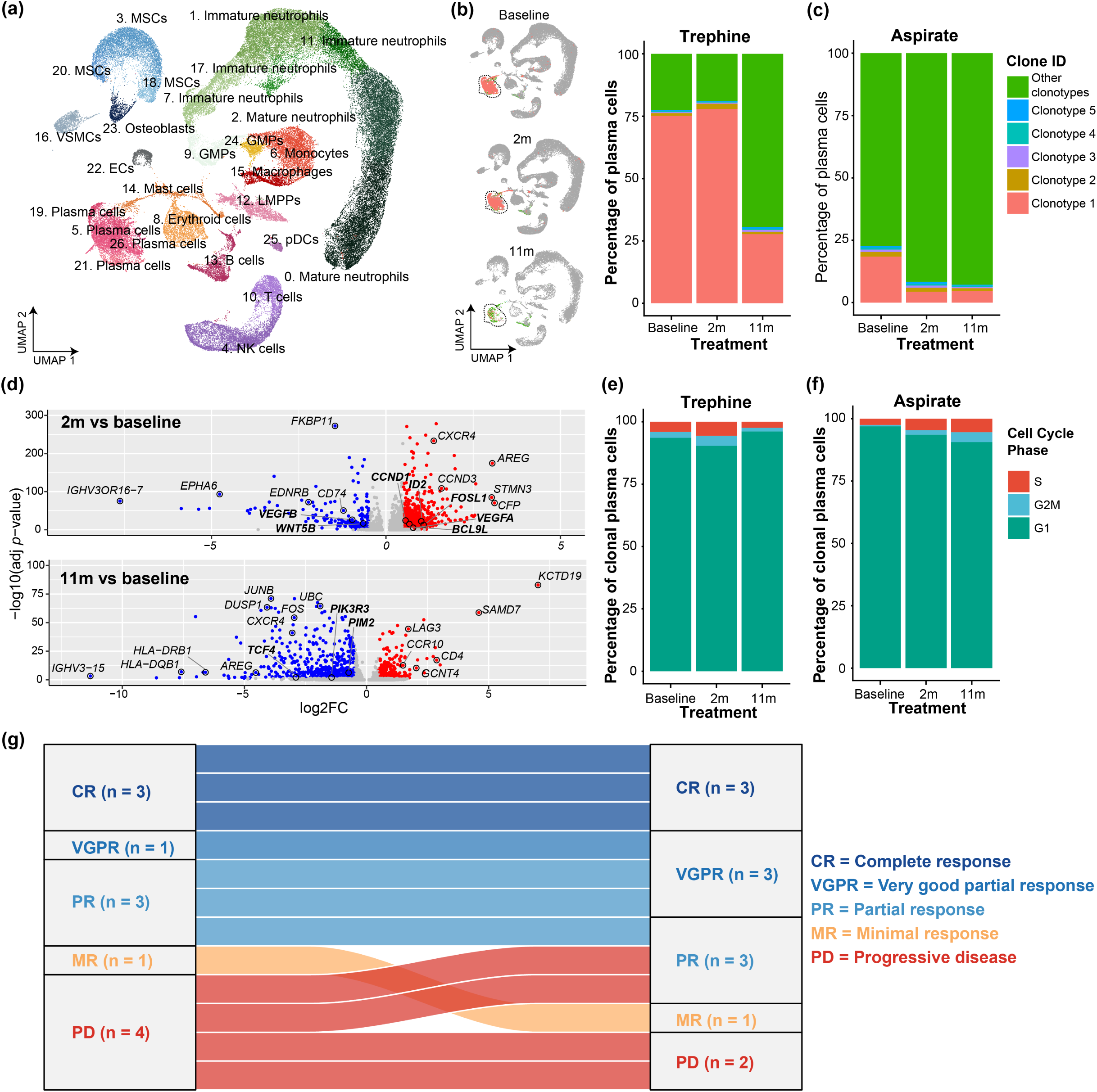
Romosozumab does not impact on myeloma disease progression **(a)** Uniform Manifold Approximation and Projection (UMAP) plot of single-cell RNA sequencing (scRNA-seq) data of cells derived from trephine biopsies, showing 27 distinct cellular clusters from all treatment timepoints. **(b)** UMAP plots and stacked bar plot showing the mapping and proportional distribution of distinct plasma cell clonotypes within human trephine and **(c)** aspirate biopsies across treatment timepoints. Dominant plasma cell clone is denoted as clonotype 1, minor clones as clonotypes 2-5 and individual cells expressing unique BCRs which are normal plasma cells are denoted as other clonotypes. **(d)** Volcano plots showing DEGs within the dominant plasma cell clone (clonotype 1) relative to baseline following 2 months and 11 months of romosozumab treatment. Genes in bold text are involved in or direct targets of Wnt pathway. Stacked bar plot showing the cell cycle phase distribution within the dominant clone in **(e)** trephine and **(f)** aspirate biopsies across all treatment timepoints. **(g)** Sankey plot illustrating the IMWG response criteria for patients at screening (left) versus end of treatment (right). Ribbon widths correspond to individual patients. For patients who discontinued or have not completed treatment yet, their post-treatment category represents that recorded at most recent folllow-up.

To assess whether romosozumab treatment was associated with altered gene expression programs in clonal plasma cells we also performed differential gene expression analysis. Two months after treatment initiation the dominant myeloma clone exhibited changes in transcriptome profiles (Fig. 4d and Suppl. Fig. 3c). This included increased expression of specific cyclins, including cyclin D1 and cyclin D3 (*CCND1* and *CCND3)* in myeloma cells from trephines (Fig. 4d) and cyclin D2 (*CCND2)* and the cyclin dependent kinase 6 (*CDK6)* in myeloma cells isolated from the marrow aspirate (Suppl. Fig. 3c). This transcriptional change was transient and returned to baseline at 11 months. Cell-cycle analysis to evaluate the proliferative dynamics of the clonal plasma cells showed that the proportion of clonal plasma cells in the actively proliferating S and G2/M phases remained unchanged across all time intervals in both bone trephine and aspirate compartments (Fig. 4e,f).

Together these data suggest that romosozumab treatment is not associated with an increase in the dominant myeloma clone or any change in clonal architecture. In support of this, all patients had the same, or saw improvements in, IMWG response criteria as was present at enrolment (Fig. 4g; Suppl. Table 2). Collectively, these longitudinal scRNA-seq and clinical data indicate that romosozumab does not exacerbate the underlying plasma cell malignancy, drive clonal expansion, or promote clinical tumour progression, supporting its safe use for the treatment of myeloma bone disease.

### Romosozumab Induces Transcriptional Reprogramming in Osteoblasts

Cells isolated and sequenced from the endosteal compartment of bone trephines included hematopoietic and non-hematopoietic cells (Fig. 4a). Analysis of hematopoietic cell clusters revealed changes in immune cell subsets in keeping with changes anticipated following chemotherapy and G-CSF regimens (Suppl. Fig. 4a). The non-hematopoietic compartment included endothelial cells (ECs), vascular smooth muscle cells (VSMCs) and cells of the osteoblast lineage, including mesenchymal stromal cell (MSC) clusters that each expressed *CXCL12* and *LEPR* (markers of *CXCL12*-abundant reticular (CAR) cells) and osteoblasts, which are the primary target for romosozumab therapy (Fig. 5a). Pseudotime analysis of predicted trajectories of the osteoblast lineage cells showed cluster 18 MSC cluster, defined by expression of polypeptide N-Acetylgalactosaminyltransferase 17 (*GALNT17)*, to be an uncommitted ‘progenitor’ population and at the start of the differentiation trajectory (Fig. 5b). Cluster 3 MSCs, expressing G-Protein Coupled Receptor 88 (*GPR88*) is an intermediate pool of cells ‘primed for differentiation’, whereas cluster 20 MSCs exhibited increased expression of adipogenic lineage and cytokine transcripts, including apolipoprotein D (*APOD)*, suggesting these are ‘adipogenic/secretory’ MSCs. Osteoblasts, which express osteocalcin (*BGLAP*), mapped to the end of the osteogenic trajectory. The proportion of osteoblasts, the cells responsible for bone formation, remained the same across the treatment timeline (Fig. 5c). There were low numbers of cells in this population, which made resolving late-stage differentiation from osteoblast progenitors and pre-osteoblasts difficult. The proportion of MSC cell populations was also similar, although *GPR88*^+^ MSCs that are primed for differentiation increased marginally, whereas *APOD*^+^ adipogenic/secretory MSCs reduced with time following romosozumab treatment (Fig. 5c). Ridgeplot analysis confirmed that romosozumab treatment did not cause significant changes in the distribution of cell populations across pseudotime (Fig. 5d). This suggests the therapeutic effect of romosozumab may be driven by effects on existing populations rather than by a significant expansion of the osteoblast population.

**Figure 5:**
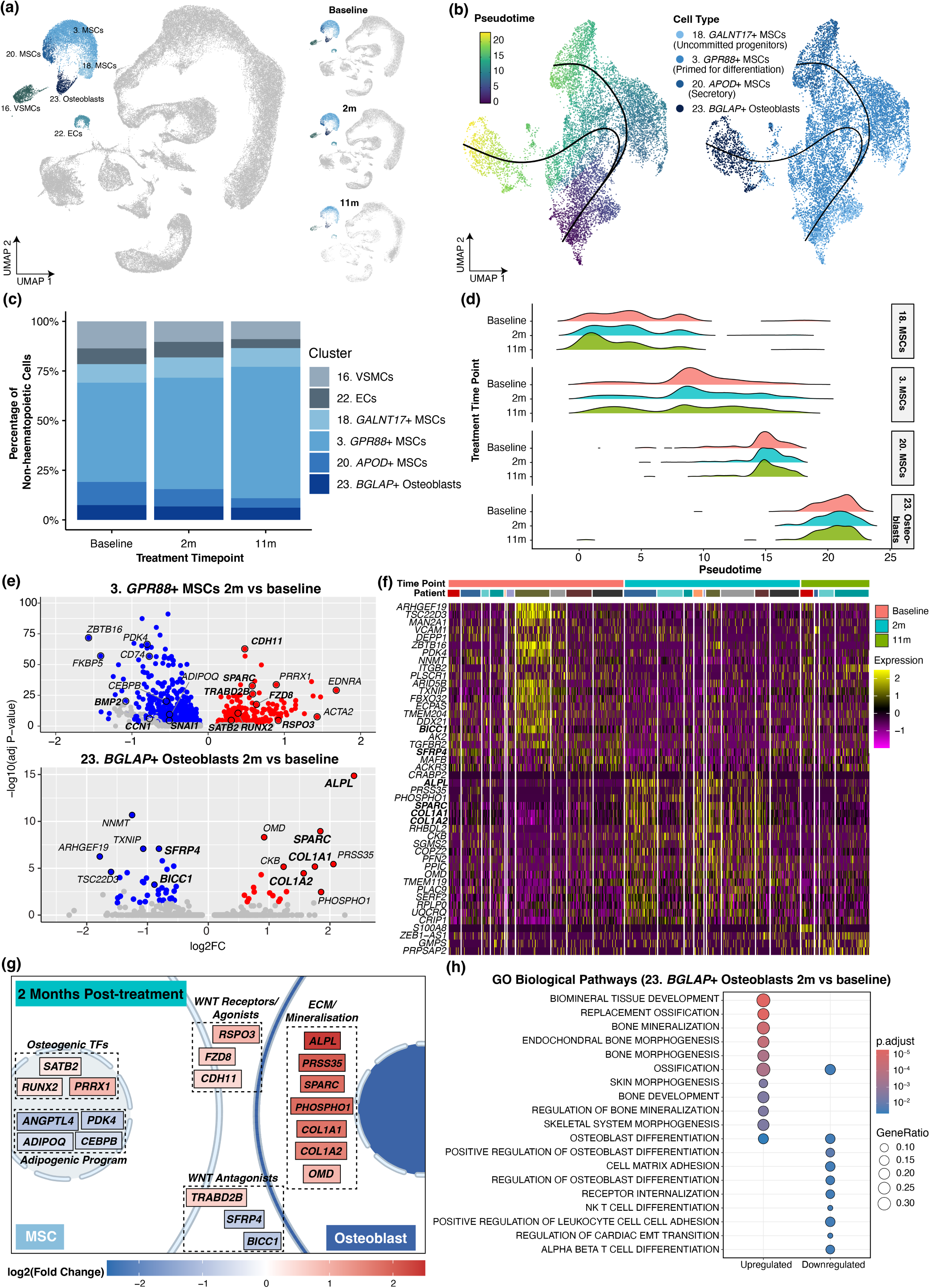
Romosozumab stimulates osteoblastic bone formation **(a)** UMAP plot of cells derived from trephine biopsies, showing the non-hematopietic cell lineages in different shades of blue and all other cells in grey from all treatment timepoints. **(b)** Differentiation trajectory analysis of mesenchymal stromal cell (MSC) and osteoblast clusters in pseudotime. **(c)** Stacked bar plot showing the proportional distribution of non-hematopoietic cell types across treatment timepoints. **(d)** Ridge plots depicting the density distribution of MSC and osteoblast clusters in pseudotime across all treatment timepoints. **(e)** Volcano plots highlighting differentially expressed genes (DEGs) at 2 months of romosozumab treatment relative to baseline within Cluster 3 *GPR88*^+^ MSCs (top) and Cluster 23 *BGLAP*^+^ osteoblasts (bottom). Genes in bold text are involved in or direct targets of Wnt pathway. **(f)** Heatmap showing patient-specific expression profiles of *BGLAP*^+^ osteoblast DEGs across treatment timepoints. **(g)** Schematic summarising coordinated transcriptional changes within MSCs and osteoblasts following 2 months of romosozumab treatment. **(h)** Gene Ontology (GO) Biological Pathway enrichment analysis of significantly upregulated and downregulated transcripts within the *BGLAP*^+^ osteoblasts at 2 months post-treatment.

To determine whether romosozumab treatment switches on gene programs that directly control bone formation we analysed differentially expressed genes in osteoblast lineage cells 2 months after treatment initiation. Both *GPR88*^+^ primed MSCs and osteoblasts exhibited significant transcriptional changes at 2 months relative to baseline, whereas *GALNT17^+^* MSC progenitors and *APOD*^+^ adipogenic/secretory MSCs remained largely unchanged (Fig. 5e; Suppl. Fig. 4b). The transcriptional changes included an upregulation of canonical Wnt pathway machinery. Frizzled 8 (*FZD8*) and R-spondin 3 (*RSPO3*), which potentiate Wnt signaling by stabilizing frizzled, were increased, whereas the Wnt antagonists soluble frizzled related protein 4 (*SFRP4*) and bicaudal C Homolog 1 (*BICC1*) were downregulated (Fig. 5e-g). Expression of the osteogenic transcriptions factors, runt-related transcription factor 2 (*RUNX2*) and paired related homeobox 1 (*PRRX1*) were increased, whereas angiopoietin-like protein 4 (*ANGPTL4*), adiponectin (*ADIPOQ*) and genes implicated in adipogenic programing were downregulated (Fig. 5e-g). As a consequence, Wnt target genes that play a critical role in bone formation, including those encoding alkaline phosphatase (*ALPL)*, collagen type 1 (*COL1A1* and *COL1A2)* and osteonectin (*SPARC)* were increased in osteoblasts, along with other genes implicated in controlling bone formation, such as osteomodulin (*OMD)* and phosphoethanolamine/ phosphocholine phosphatase 1 (*PHOSPHO1),* which is important in mineralisation (Fig. 5e-g). Gene Ontology (GO) analysis of differentially expressed genes in osteoblasts revealed significant enrichment of gene programs associated with biomineral tissue development, bone mineralisation and ossification (Fig. 5h). This was accompanied by a down-regulation of gene programs involved in early differentiation. Together these data suggest that romosozumab induces the synchronised induction of Wnt signaling across the osteogenic lineage in a coordinated manner that culminates in a switch to matrix synthesis and mineral deposition in osteoblasts.

In osteoblasts, the changes in gene programs, including those associated with matrix synthesis and mineralisation, returned to baseline at 11 months (Suppl. Fig. 4c), suggesting the anabolic window had closed. *APOD*^+^ MSCs also showed a reduction in differential expression (Suppl. Fig. 4d). In contrast, *GPR88*^+^ MSCs and *GALNT17*^+^ MSCs had sustained transcriptomic activity. In *GPR88*^+^ MSCs expression pivoted towards programs that would put a brake on Wnt signalling (Suppl Fig. 4e). This included increases in TRAB domain-containing 2B (*TRABD2B),* a membrane metalloproteinase that cleaves Wnt ligands; glycogen synthase kinase-3 beta (*GSK3B*); and the transcription factor 7 like 1 (*TCF7L1*), a repressor of Wnt signalling. This was associated with a down-regulation of β-catenin (*CTNNB1*), the central mediator of canonical Wnt signaling. This inhibition of Wnt signaling was accompanied by a downregulation of immediate-early response/AP-1 transcription factors (*FOS*, *FOSB*, *JUN*, *JUNB*, *EGR1*), reflecting a shutdown of mitogenic signals, as well as a reduction in insulin-like growth factor 1 and 2 (*IGF1* and *IGF2*) and IGF binding proteins (*IGFBP3*, *IGFBP5*). Collectively, these data demonstrate that by 11 months, MSCs in the endosteal compartment deploy cell-autonomous feedback machinery to silence canonical Wnt and IGF signaling. Together this serves to limit osteoblast bone formation and return levels to baseline.

## DISCUSSION

Myeloma bone disease causes significant morbidity and mortality^3^. A significant proportion of patients continue to lose bone, develop new lytic lesions and sustain fractures while receiving zoledronic acid or denosumab, the current standards of care. The field has come to accept this residual burden of skeletal morbidity. The theoretical concerns that pharmacological activation of Wnt signalling could stimulate myeloma cell proliferation have meant that Wnt-targeted strategies that restore lost bone and have transformed outcomes in osteoporosis have not been explored in patients with cancer. Patients who are refractory to antiresorptive therapy therefore have no meaningful treatments to stop further bone loss or rebuild lost bone and strengthen their skeleton. The data in this study address this gap and these safety concerns.

Romosozumab was well tolerated, with grade 1-2 injection site reactions being the only TEAE. Notably there were no reports of hypocalcaemia, osteonecrosis of the jaw or cardiovascular events. This was reassuring given this cohort was unselected and included patients with active myeloma. Among the patients with sequential bone marrow samples available, single-cell transcriptional profiling of the malignant plasma cell compartment showed no expansion of the myeloma clone, no change in clonal architecture, and no transcriptional signature associated with accelerated proliferation during treatment. This is consistent with preclinical data that demonstrated no increase in myeloma burden and no reduction in survival following treatment. Clinically, no patient on study progressed through an IMWG response criteria. Two patients who were enrolled with progressive disease continued to progress on study (one new, asymptomatic lesion identified on PET-CT and one treatment refractory disease leading to palliation). Of note, in a prior observational study of 8 post-menopausal women with osteoporosis and smouldering myeloma treated with romosuzumab, no patient progressed to symptomatic myeloma^40^. Together, these prospective data show sclerostin inhibition is safe in myeloma, removing a critical barrier to the development of Wnt-targeted bone anabolic therapies for cancer patients.

Romosozumab treatment produced a rapid and substantial rise in PINP, a serum marker of bone formation. Treatment had no effect on beta-CTX, a marker of bone resorption which likely reflects the pre-treatment with zoledronic acid. Single-cell analysis at 3 months showed the proportion and differentiation trajectory of mesenchymal stromal cells and osteoblasts to be unchanged. However, the transcriptional state of osteoblasts transitioned to active bone formation, with upregulation of gene programmes governing collagen synthesis, mineralisation, and overall osteoblast function. The anabolic effect of romosozumab appears to be driven principally by reprogramming of existing committed osteoblasts rather than by recruitment of new progenitors. Bone formation markers declined by 12 months but remained above baseline and higher than reported in the pivotal osteoporosis trials. This likely reflects the distinct biology of the myeloma microenvironment rather than prior zoledronic acid exposure, since romosozumab acts predominantly through modelling-based bone formation on quiescent surfaces rather than remodelling-based formation coupled to prior resorption^41–43^.

The increases in bone formation translated into clinically meaningful increases in bone. Whole-body Hounsfield units measured by CT and BMD measured by DXA increased substantially over 12 months. Increases in bone were observed across all anatomical sites. This is important given both generalised bone loss and focal bone lesions contribute to myeloma bone disease and fractures are not restricted to the axial or appendicular skeleton. At the lumbar spine and proximal femur, the magnitude of BMD increase was comparable to, and in many cases exceeded, those seen in the pivotal osteoporosis studies^26,32^. This is despite our cohort comprising patients with established malignant bone disease who had received prior, often prolonged, antiresorptive therapy. High-resolution three-dimensional analysis of the different bone compartments showed increases were not restricted to the trabecular compartment but extended to cortical bone, a finding replicated in our preclinical models. This carries particular importance, as cortical thickness and porosity are major determinants of bone strength and resistance to fracture. Critically, the combined trabecular and cortical response offers a mechanistic rationale to expect improvements in bone density to translate into increases in bone strength and fracture resistance.

Beyond global gains in bone density, romosozumab also partially repaired pre-existing osteolytic lesions in a subset of patients. Cross-sectional CT analysis showed that this radiological response was mediated by the deposition of new bone onto existing bone surfaces bordering each lesion, rather than de novo bone formation across the entire lesion. Not all lesions responded, and those that did remained radiologically detectable at the end of treatment. This likely reflects the volume of bone needed to refill lesions created by years of unopposed osteoclast activity rather than therapeutic failure. Extending lesion repair will require rethinking how bone anabolic agents are deployed in the myeloma treatment landscape. Whilst antiresorptives remain the current frontline standard-of-care in myeloma, recent paradigm shifts in the osteoporosis setting demonstrate that an “anabolic-first” strategy of initiating romosozumab and consolidating with an anti-resorptive, yields superior effects than when used as a salvage therapy^29,30,44–46^. Extrapolating this ‘anabolic-first’ strategy to multiple myeloma, romosozumab may have a greater clinical impact to rapidly rebuild skeletal integrity when used de novo and followed by standard zoledronic acid to consolidate and maintain the newly formed bone. Defining the optimal strategy is both an opportunity and a clear priority for future trials.

The single-cell data also offer a tractable approach to understanding the biphasic response to sclerostin inhibition. At three months we observed a coherent anabolic transcriptional response. Local Wnt signalling was amplified through induction of *FZD8* and the agonist *RSPO3* and suppression of *SFRP4* in osteoblasts. The osteogenic transcription factors *RUNX2* are upregulated, adipogenic fate programs were actively repressed and osteoblasts engaged robust matrix-synthesis and mineralisation gene programs. By twelve months this gene signature was down-regulated despite continued sclerostin inhibition. The brake appeared to be located inside the cell at the level of the destruction complex and transcriptional control rather than at the level of romosozumab binding sclerostin. β-catenin was downregulated, *GSK3B* was upregulated, and the Wnt-target repressor *TCF7L1* was induced. This was accompanied by down regulation of AP-1 / immediate-early genes and MSC-derived IGF1 and IGF2. What initiates this intracellular re-programming remains unclear. The osteoprogenitor pool could be exhausted following sustained anabolic demand, alternatively the MSC/osteoblast lineage cells could undergo epigenetic reprogramming locking cells in an insensitive state. Metabolic changes may also contribute as Wnt signalling regulates the glycolysis-to-oxidative phosphorylation hand-off during osteoblast differentiation. Understanding these pathways will be important in identifying combination strategies that could sustain the bone anabolic effect of romosozumab.

Several limitations should be acknowledged. This was a phase IIa proof-of-concept study designed to establish safety, tolerability, and biological proof of concept, and the cohort was modest in size and heterogeneous with respect to disease status, prior and ongoing myeloma treatment. All patients received romosozumab on a background of prior zoledronic acid, and the impact of romosozumab in the absence of previous antiresorptive therapy cannot be inferred. The single-cell mechanistic analyses were evaluated at two– and 11-month timepoints. Transient or rapidly resolving effects on mesenchymal stromal cell differentiation occurring early may not have been captured. Osteocytes and osteoclasts are not sampled using this methodology so impacts on sclerostin (*SOST*) or dickkopf-1 (*DKK1)* were not measured. Whether the molecular understanding is replicated at a protein level is also unknown. Finally, while no patient experienced a new SRE that was clearly attributable to disease progression during treatment, follow-up beyond the 12-month treatment window will be required to define the durability of bone gains and impact on new SREs.

Together this study provides prospective clinical evidence that sclerostin neutralisation with romosozumab is safe, well tolerated, and biologically active in patients with multiple myeloma who continue to experience skeletal-related events in bone despite standard antiresorptive therapy. Romosozumab stimulated osteoblast function, increased bone formation markers and produced substantial gains in trabecular and cortical BMD. We saw evidence of repair to pre-existing osteolytic lesions alongside a profound reduction in SREs. Romosozumab did so without evidence of myeloma progression at the clinical, clonal, or transcriptional level. These findings provide the mechanistic and clinical rationale to evaluate sclerostin inhibition in adequately powered, randomised controlled trials. These data combined with the movement in the osteoporosis field towards an “anabolic-first” strategy, provide the rationale for randomised studies of romosozumab upfront followed by zoledronic acid in newly diagnosed multiple myeloma. As myeloma patients continue to benefit from novel targeted therapies, these data offer a vital, parallel step towards actively rebuilding the skeleton ensuring that extended overall survival is coupled with restoration of skeletal health.

## METHODS

### Animal experiments

Animal experiments were approved by the Garvan Institute of Medical Research Animal Ethics Committee (ARA22/12, ARA25/23). C57BL/KaLwRij mice (Harlan, The Netherlands) were injected intravenously with syngeneic 5TGM1-eGFP or saline. Mice were injected, subcutaneously, once per week, with romosozumab (MedChemExpress, 10mg/kg) or vehicle, commencing on day 1 for up to 26 days.

### Flow cytometry analysis of myeloma burden in mice

Cells were isolated from bone marrow of right femur and spleen. Isolated cells were incubated with red blood cell lysis buffer before staining with DAPI (Sigma-Merck) and analysed on a BD Symphony analyser (BD Biosciences).

### Estimated paralysis degree (EPD) scoring of myeloma-bearing mice

Hindlimb paralysis was evaluated using the Experimental Paralysis Degree (EPD) scoring method, an adaptation of the Experimental Autoimmune Encephalomyelitis (EAE) scoring system^47^. To ensure objectivity, a blinded daily assessment of each animal’s motor function, reflexes, and overall clinical condition was performed. Mice were scored according to the following standardised criteria:

0.0: Normal behavior, no clinical signs of disease.

0.5: Partial loss of tail tonicity or flaccidity of the distal tail region, occasionally accompanied by a hunched appearance.

1.0: Completely limp tail with no curling at the tip.

1.5: Complete tail paralysis combined with distinct weakness in the hind limbs.

2.0: Waddling gait, weakness in the hind limbs, or dragging of one hind limb (unilateral partial hindlimb paralysis).

2.5: Complete dragging or partial paralysis of both hind limbs (bilateral).

3.0: Complete paralysis of both hind limbs. The mouse may drag its hindquarters but can still move its forelimbs forward.

3.5: Complete hind limb paralysis accompanied by partial paralysis or weakness in one forelimb.

4.0: Quadriplegia (complete paralysis of all four limbs) or severe paralysis of hind limbs and weakness of front legs.

5.0: Moribund state or dead

### Measurement of PINP and TRAP in mouse serum by ELISA

Serum collected by retro-orbital bleeds at endpoint under anesthesia with isoflurane, was stored at −80°C and then assessed for PINP and TRAP5b levels using ELISA kits (Immunodiagnostic Systems) following the manufacturer’s instructions.

### DXA analysis of bone mineral density in mice

DXA was performed using a Faxitron UltraFocus (Hologic) on mice anesthetised under 3%-5% inhaled isoflurane on day 0, day 14 and endpoint. Hindlimb analysis was performed using VisionDXA (Hologic) and manually drawn regions-of-interest (ROI) encompassing the hindlimb excluding the foot, and L2 vertebra, were used to quantify BMD.

### MicroCT analysis of mouse femur and vertebrae

Formalin-fixed left femora and the L2 vertebrae were imaged with a SkyScan 1172 microCT scanner (Bruker) at a resolution of 4.3 μm, 0.5 mm aluminum filter, 50 kv voltage, and 200 μA current. Images were captured every 0.4° through 180° and were reconstructed and analyzed using NRecon software (SkyScan). Bone structural parameters and nomenclature were utilised according to standardised guidelines. Three-dimensional reconstructed images of femora were generated using Drishti imaging software version 2.4 (ANU). ROI selection and analysis were performed using CTAn software (Bruker). To examine the changes in femoral bone parameters trabecular bone parameters were calculated from scans performed at a voxel resolution of 4.4 μm in a 1.5 mm region beginning 500 μm proximal to the distal femoral growth plate to reduce the contribution of the primary spongiosa in the analysis. Cortical bone parameters were calculated from scans performed at a voxel resolution of 4.4 μm in a 1 mm region mid diaphysis. Changes in vertebral bone trabecular and cortical parameters were calculated from scans performed at a voxel resolution of 4.4 μm in an ROI calculated by measuring the distance between 0.2 mm offset from the point of 50% spongiosa and trabecular bone on both ends of the vertebral body.

### Statistical methods for animal experiments

Statistical analyses were performed using GraphPad Prism (v9.0; GraphPad Software). For multi-group comparisons, a one-way ANOVA with Tukey’s post-hoc correction was applied. Longitudinal or grouped datasets were evaluated via a mixed-effects analysis or two-way ANOVA with Šídák’s multiple comparisons test. Differences between two independent populations were assessed using an unpaired t-test. All data are expressed as mean ± standard deviation (SD). Statistical significance was defined as *p* < 0.05.

### Clinical Trial design

This phase II, single-arm, proof-of-concept trial investigated the safety and efficacy of romosozumab in patients with multiple myeloma. The trial was a collaborative initiative between St Vincent’s Hospital, Sydney, and the Garvan Institute of Medical Research (ANZ trials registry ACTRN12624000027516). Patients were assigned to receive subcutaneous injections of romosozumab (at a dose of 210 mg) once monthly for 12 months. All existing anti-resorptive treatments, such as zoledronic acid, were ceased upon study enrolment. Throughout the treatment phase, all patients were additionally prescribed daily calcium (500–1000 mg) and vitamin D (500–1000 IU) supplementation.

### Trial oversight

The trial protocol and informed consent documents were approved by the Human Research Ethics Committee (HREC) of St Vincent’s Hospital, Sydney. The study was performed in strict accordance with the National Health and Medical Research Council (NHMRC) Statement on Ethical Conduct in Human Research, and the principles laid down in the Declaration of Helsinki. All patients provided written informed consent prior to the commencement of any trial-specific procedures.

### Patients

Ambulatory adult patients (aged 18 years or older) with a confirmed diagnosis of multiple myeloma were eligible for participation if they had experienced a skeletal-related event despite receiving at least three doses of prior anti-resorptive therapy. Patients were excluded if they had a history of myocardial infarction or stroke within the previous 12 months, active unstable cardiovascular function (including a left ventricular ejection fraction below 50% or congestive heart failure of New York Heart Association Class 3 or higher), uncorrected vitamin D insufficiency (levels < 50 nmol/L), or a history of osteonecrosis of the jaw. Women of childbearing potential and patients previously treated with teriparatide or parathyroid hormone (PTH) analogues were also excluded.

### Procedures

Fasted serum concentrations of the bone-turnover markers procollagen type 1 N-terminal propeptide (PINP) and the beta-isomer of C-terminal telopeptide of type I collagen (beta-CTX) were measured at baseline, and at months 1, 2, 3, 6, and 11. Bone mineral density (BMD) at the total hip was evaluated via dual-energy x-ray absorptiometry (DXA) at baseline, 6 months, and 12 months. Computed tomography (CT) skeletal surveys were obtained at baseline, 12 months, and 24 months. These were retrospectively reviewed along with clinical notes and reports from previous accessible imaging to determine the number of lesions existing at the time of study commencement and the number of SREs encountered since diagnosis.

Patients’ myeloma disease was monitored by serum protein electrophoresis, immunofixation and serum free light chains at baseline and at months 2, 5, 8 and 11. In addition, referring physicians were asked to communicate any treatment changes or clinical disease progression.

Additionally, paired bone marrow aspirates and bone trephines were collected at baseline, between the second and fourth doses, and at 11 months.

### Outcomes

The primary end points of this trial were the safety of Romosozumab, determined by adverse events graded according to the Common Terminology Criteria for Adverse Events (CTCAE) version 5, and the effect of Romosozumab treatment on the bone turnover marker PINP. Secondary end points included the percentage change from baseline in lumbar spine and total hip bone mineral density at 6 and 12 months and the incidence of SREs (defined as new fractures, or the need for radiotherapy or surgery) at 3, 6, and 12 months. Exploratory objectives included determining radiological improvements in bone lesions on CT imaging; percentage change in Int_vBMD, Trab_vBMD, Cort_vBMD, sBMD and Cort_Thick at 6 and 12 months relative to baseline; HU across individual lesions and throughout the whole body; and assessing dynamic cellular changes in the bone marrow microenvironment via RNA sequencing.

### Exploratory imaging analysis

#### DXA-based 3D modelling

To further evaluate trabecular and cortical compartments, hip DXA scans taken at baseline, 6 and 12 months were analysed using 3D modelling software (3D-SHAPER v2.14.0, 3D SHAPER MEDICAL) to calculate integral, trabecular and cortical volumetric BMD (Int_vBMD, Trab_vBMD, Cort_vBMD) (g/cm3), cortical thickness (Cort_Thick mm) and cortical surface BMD (sBMD) (g/cm2) as previously described^34,35^.

#### Assessment of size and radiodensity across lytic lesions

Retrospective review of CT imaging (non-contrast CT skeletal survey extending from the calvarium to just below the knees) was performed by a Board-Certified Radiologist for all patients that had paired pre and post treatment imaging (n=7). One patient did not have a pre-treatment skeletal survey and the low dose attenuation correction CT component of a PET-CT was substituted.

Imaging review and measurements were performed on Phillips Vue PACS version 12.2.8.450.0001. For each patient, pre and post treatment linear and Hounsfield unit measurements were obtained of the largest identifiable and measurable myelomatous lesion, defined as a well-circumscribed lytic lesion. Linear measurements were obtained in orthogonal planes, measured in millimetres. Hounsfield unit measurements were obtained by measuring the attenuation of the largest lytic area within a lesion. Following IMWG methodology^33^, the sum of the products of cross-diameters was used to determine lesion size. One lesion could only be measured in a single dimension and therefore was squared for relativity to the others. This lesion is highlighted as 1D in figure 3b.

To obtain radiodensity data (Hounsfield Units (HU)) across lesions and graphically illustrate radiologically visible bone formation along the periphery of a lesion on post-treatment images, a line profile extending across the lesion along the plane of visible bone formation (if present) was drawn in the open-source software 3D Slicer (version 5.10.0). A shorter line along the same plane was obtained on the same software to demarcate the radiological limit of lesion extent. Similar line profiles were obtained on lesions without visible change with measurements taken in the longitudinal or short axis of a lesion. HU across each line was extracted by loading the original DICOM series and reconstructing a 3D volume. The line endpoints were parsed from 3D Slicer’s JSON markup files, and HU were sampled at 400 equispaced points between the endpoints using trilinear interpolation. The shorter line was used to mark the lesion boundaries on the profile. Pre– and post-treatment lines were oriented in the same direction so that the two profiles could be plotted on a common distance axis for comparison.

#### AI-assisted bone segmentation

To expand our DXA-generated areal BMD data, CT skeletal survey images for all patients with paired pre– and post-treatment scans (n=7) were processed using Skellytour (medium model), a deep learning segmentation framework built on nnU-Net^37,38^. Skellytour was trained on datasets that include osteolytic lesions and MM-related skeletal deformity, reducing the risk of segmentation failure at sites of active disease. The medium model produces 38 anatomical labels: individual vertebrae (C1–L5), paired labels for femora, ribs, humeri, scapulae, and clavicles, and combined labels for skull, pelvis, and sternum.

A whole-body median radiodensity was calculated across all segmented bone voxels (background and the artifact label excluded). In addition, median HU data were extracted from seven segmented regions of interest; cervical, thoracic, and lumbar vertebrae, bilateral ribs, and bilateral femora. The humerus was prospectively excluded from comparative analyses due to inconsistent field-of-view truncation between pre– and post-treatment scans.

### Statistical analysis of trial outcomes and exploratory analysis

The pilot study enrolled a target sample size of 12 participants. No hypothesis testing was performed in advance due to the lack of data available to calculate estimated effect size and the primary objective being safety.

Longitudinal changes in bone mineral density (DXA and 3D shaper data) and bone turnover markers (PINP and beta-CTX) were evaluated using linear mixed-effects models (LMM) with restricted maximum likelihood (REML) estimation. To account for missing data and the repeated-measures design, patient identification was included as a random intercept allowing each patient to serve as their own baseline control adjusting for time-invariant between-subject characteristics. Timepoint was treated as a categorical fixed effect. Degrees of freedom and standard errors of the adjusted mean were calculated using the Satterthwaite approximation. The main effect of time was assessed via Type III Analysis of Variance (ANOVA), with specific baseline comparisons extracted from model coefficients. For graphical visualization, Estimated Marginal Means (EMMs) were extracted and plotted as percentage changes from baseline alongside raw individual patient data points and cohort medians. Despite this being a phase II pilot study, Dunnett’s adjustment was applied to all p values related to multiple comparisons. All statistical analyses were performed using R software (version 4.6.0 (2026-04-24 ucrt), utilizing the lmerTest and emmeans packages. Statistical significance was defined as a two-tailed p-value < 0.05.

CT segmentation data analysis was performed using Python (version 3.12). For each patient, a median HU was calculated per skeletal region. For paired or regional skeletal groups (e.g. bilateral femora or vertebrae), a single aggregated patient-level value was generated by calculating the median of the constituent sub-regions. Non-parametric statistical tests were used given the small sample size and exploratory nature of this sub-study. Changes in pre– and post-treatment scans were calculated using the exact, one-sided Wilcoxon signed-rank test. Effect sizes were calculated using the matched-pairs rank-biserial correlation (r). Descriptive statistics for the CT data are reported as the median and interquartile range (IQR).

### Cell isolation from trephine bone biopsies and marrow aspirates

Trephine biopsies and marrow aspirates were obtained from patients at 2 months and 11 months after the start of romosozumab treatment. Trephines were cut into smaller pieces using surgical blade and incubated in PBS containing 2mg/ml of collagenase A (Merck) and 2.5mg/ml of dispase (Merck) for 30 mins at 37°C. After digestion, trephine samples were vortexed for 10s and the supernatant containing digested cells was filtered through a 100μm filter into collection tubes containing 10% fetal calf serum (FCS; Bovogen Biologicals). Cells were then incubated in red blood cell lysis buffer for 5 mins at RT (Roche). Marrow aspirate cells were processed by centrifugation and incubated with red blood cell lysis buffer for 5 mins at RT.

### FACS enrichment of cells

Trephine cells were stained for CD235a-BUV395 (BD Biosciences, 563810) and CD45-APC-H7 (BD Biosciences, 641408) at 4°C for 30 mins and rinsed with PBS supplemented with 2% FCS. Dead cells and debris were excluded by FSC, SSC and DAPI (ThermoFisher Scientific). Cells that were viable (DAPI-negative) and negative for erythroid marker (CD235a) were then sorted based on CD45 for haematopoietic (CD45-positive) and non-haematopoietic (CD45-negative) cells into PBS supplemented with 2% FCS.

Aspirate cells were stained for CD235a-BUV395, CD38-AF488 (BioLegend, 356634) and CD138-BV785 (BioLegend, 356538) at 4°C for 30 mins and rinsed with PBS supplemented with 2% FCS. Dead cells and debris were excluded by FSC, SSC and DAPI. Cells that were viable (DAPI-negative) and negative for erythroid marker (CD235a) were then sorted based on CD38 and CD138 (plasma cells) into PBS supplemented with 2% FCS.

### Single cell RNA-seq (scRNA-seq) and data analysis

Cells were encapsulated into emulsion droplets using the 10x Chromium platform (10x Genomics). scRNA-seq libraries were constructed using the Chromium GEM-X Single Cell 5’ B-Cell Receptor (BCR) Reagent Kit according to the manufacturer’s protocol. Briefly, FACS sorted cells were examined under a microscope and counted with a cell counter (Thermo Fisher Scientific). Cells were loaded into each channel with a target output of 10,000 cells. Reverse transcription and library preparation were performed on a C1000 Touch Thermal cycler with 96-Deep Well Reaction Module (Bio-Rad). Amplified cDNA and final libraries were evaluated on an Agilent Tapestation using a High Sensitivity D1000 ScreenTape (Agilent Technologies, 5067-5584). Individual libraries were diluted to 4nM and pooled for sequencing. Pools were sequenced with 75 cycle run kits (26bp Read1, 8bp Index1 and 55bp Read2) on the Novaseq Sequencing System (Illumina) to 80-90% saturation level. scRNA-seq services were provided by the Garvan Genomics Platform at the Garvan Institute of Medical Research.

Raw data were processed with CellRanger (v7) and analysed using the Seurat package (v5)^48^. Log normalisation was performed and low-quality cells filtered (genes<600 or Mitochondrial UMI>15-20%). Dimensionality reduction used PCA (20 PCs) on the top 2000 variable genes. Data was batch-corrected using Harmony^49^. Clustering used the Louvain Method on shared nearest neighbour graphs and visualised by UMAP. Cell types were annotated using canonical markers and cluster-specific gene programs, as determined by Seurat FindAllMarkers, requiring log2FC>0.5 and *Bonferroni*-adjusted *P*<0.05. A cluster was deemed to express a gene if 1 or more UMI for that gene was detected in at least 10% of cells within the cluster.

Differential gene expression across specific clusters at 2 and 11 months versus baseline was evaluated using Seurat FindMarkers. Significant differentially expressed transcripts were defined by a *Bonferroni*-corrected and a minimum detection rate of 10% of cells within the respective cluster.

### Reconstruction of cell differentiation trajectories

To reconstruct differentiation trajectories, lineage inference was performed using the Slingshot package (v 2.18.0)^50^. Unsupervised cluster assignments and dimensionality reduction coordinates (UMAP/PCs) were utilised as inputs. A minimum spanning tree was constructed across the clusters to establish branching lineages, followed by the fitting of simultaneous principal curves to model smooth cellular trajectories. Pseudotime values were subsequently assigned to individual cells based on their orthogonal projections along these curves.

### Single-Cell BCR Clonal Analysis

Cells were assigned to distinct clonotypes based on the strictly paired tracking of identical nucleotide sequences for both productive heavy– and light-chain V(D)J junctions. Plasma cells containing incomplete or non-productive pairings were filtered out of the clonal frequency calculations. The dominant clone was identified based on its proportional mass expansion relative to the total captured plasma cell population within each tissue compartment. Clonal tracking data were subsequently integrated with the single-cell RNA-seq expression matrices to analyze clonotype-specific transcriptomic changes across individual treatment timepoints.

## DATA AVAILABILITY

All data will be made available upon reasonable request. All raw scRNA-seq datasets will be made publicly available via the Gene Expression Omnibus (GEO).

## CODE AVAILABILITY

All single-cell transcriptomic analyses and statistical analyses of clinical data were performed using publicly available software as described in Methods. Custom scripts will be made publicly available via GitHub and Zenodo.

## AUTHOR’S CONTRIBUTIONS

P.I.C., G.M., J.M., and T.G.P., conceived the study. R.C.C., B.G., P.I.C. and G.M., designed experiments. R.C.C., A. C-M., E.S., S.G.Y, C.M.S and J.T.S., performed mouse experiments. G.M, B.G., A.B., J.C. and P.I.C designed the clinical study. B.G., G.M., J.P., L.L. and S.L., recruited patients and collected samples. B.G., N.P., X.L., C.Y. and J.C., undertook clinical imaging and analysis of imaging data. R.C.C., S.G.Y, E.S., and J.T.S. isolated bone marrow and bone trephine cells and performed scRNA-seq analysis. B.G., R.C.C., C.L.C., K.N.W., J.M., T.G.P., P.I.C., and G.M., analysed and interpreted data. R.C.C., B.G., G.M., and P.I.C., wrote the manuscript. All authors reviewed and revised the manuscript.

## ACKNOWLEDGEMENTS

This work was supported by Tour de Cure, Blood Cancer United, Cancer Council NSW, Mrs. Janice Gibson and the Ernest Heine Family Foundation. P.I.C., T.G.P. and C.L.C., are supported by NHMRC Investigator Grants (GNT2009010, GNT1155678, GNT2033083). J.T.S. was funded by a UNSW Sydney International Postgraduate Award. Additional support came from the UNSW Cellular Genomics Futures Institute and a National Breast Cancer Foundation Collaborative Research Accelerator grant (2024/CRA0020).

We thank Alice Powell, Kimberley Razay, Alyssa Pantalone, Joshua Neish, Susan Stapleton, Catherine Lancaster and Michelle Ward for their dedication to patient care. We thank Suat Dervish, Eric Lam, Yasmin Husaini, Chris O’Keefe, Alejandro Rios Villamil, Elizabeth Tay, and the staff of the Garvan Genomics Platform at the Garvan Institute of Medical Research for support with FACS and scRNA-seq. We also thank the Data Science Platform at the Garvan Institute for support with high-performance computing, and the Australian BioResource Biological Testing Facility.

## TABLES

**Supplementary Table 1:** MicroCT data from romosozumab-treated mice.

**Supplementary Table 2:** IMWG disease classifications pre and post treatment.

| Patient | IMWG disease classification |  |
| --- | --- | --- |
|  | Baseline | End of treatment period |
| 1 | Progressive disease | Partial response |
| 2 | Progressive disease | Progressive disease |
| 3 | Partial response | Very good partial response |
| 4 | Minimal response | Minimal response |
| 5 | Complete response | Complete response |
| 6 | Complete response | Complete response |
| 7 | Partial response | Very good partial response |
| 8 | Progressive disease | Partial response |
| 9 | Complete response | Complete response |
| 10 | Very good partial response | Very good partial response* |
| 11 | Progressive disease | Progressive disease* |
| 12 | Partial response | Partial response* |

\*Remain on treatment.

## FIGURE LEGENDS

**Supplementary Figure 1:**
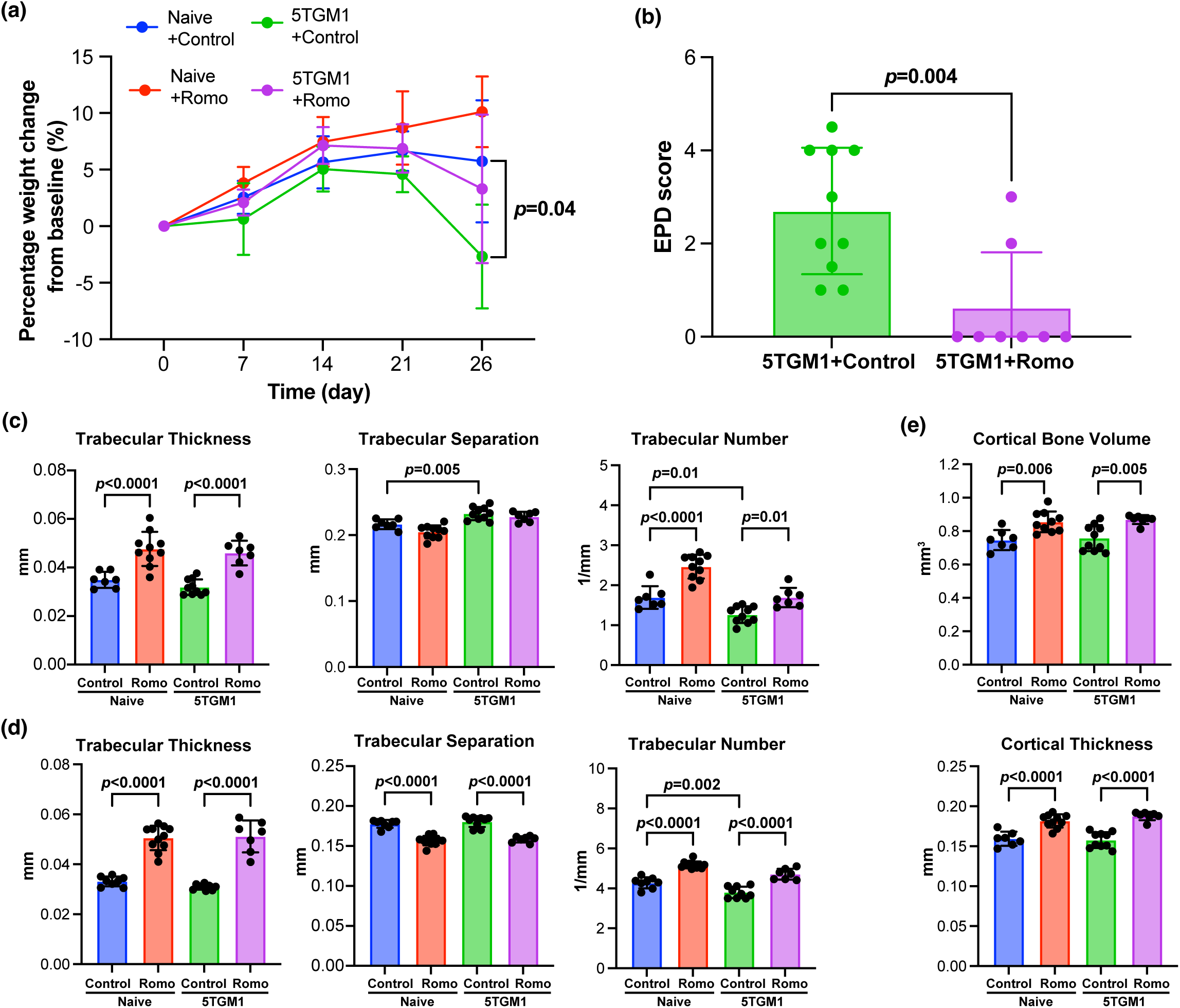
Romosozumab improved morbidity and bone mass without affecting tumour growth in murine model of myeloma **(a)** Longitudinal body weight expressed as the percentage change from baseline across all treatment groups. **(b)** End-point Experimental Paralysis Degree (EPD) scoring utilised as an indication of animal morbidity. MicroCT quantification of **(c)** distal femoral and **(d)** L2 vertebral trabecular parameters, including thickness (Tb.Th), separation (Tb.Sp), number (Tb.N), and **(e)** femoral mid-shaft cortical parameters, including cortical bone volume (BV) and thickness (Ct.Th). Data are presented as mean ± SD. Statistical significance was evaluated using a mixed-effects model with Tukey’s multiple comparisons test for **(a)** and an unpaired Welch’s *t*-test for **(b)** and one-way ANOVA with Tukey’s multiple comparisons test for all other parameters.

**Supplementary Figure 2:**
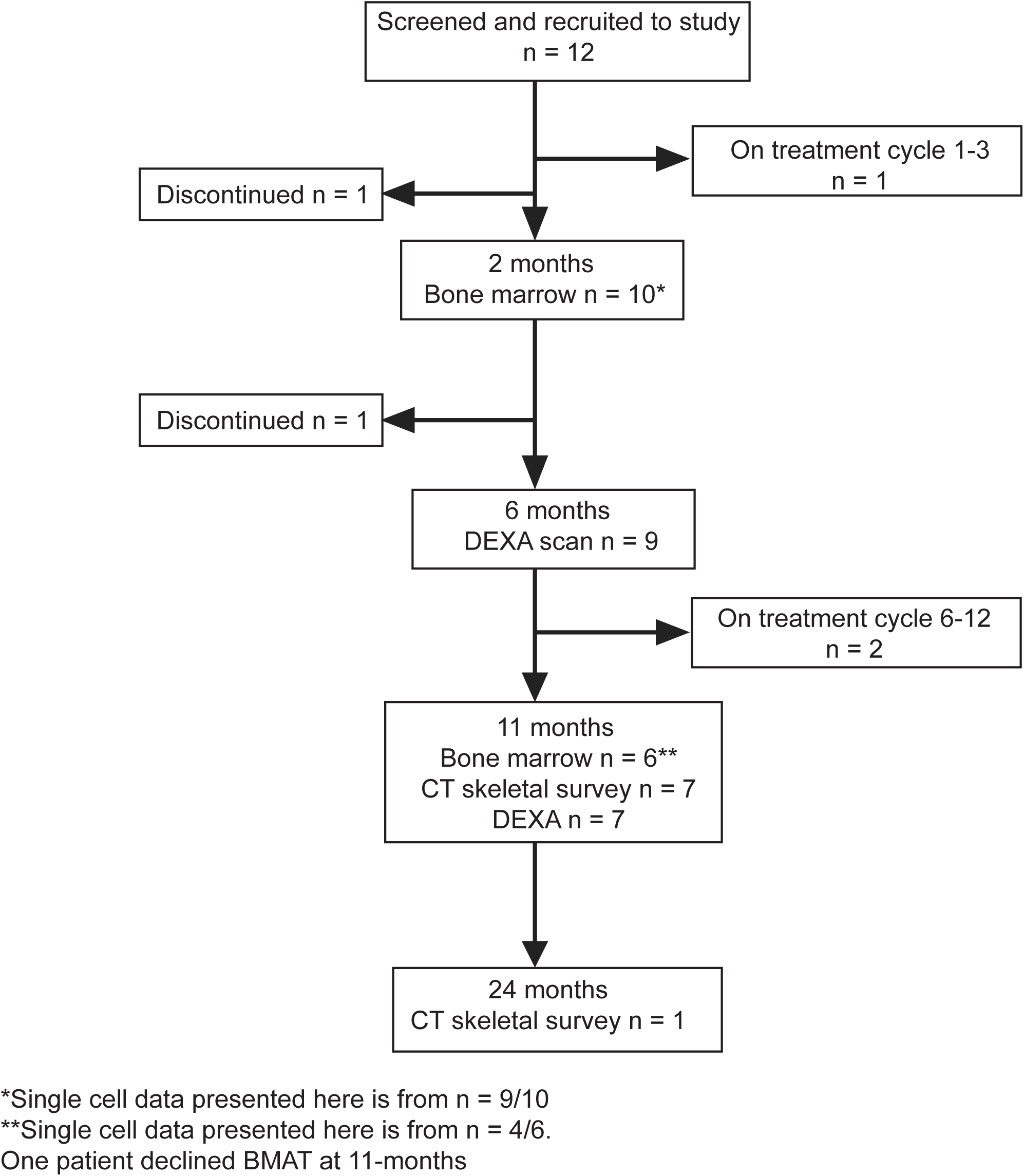
Consort diagram of patient enrolment and progress through the study as of this publication.

**Supplementary Figure 3:**
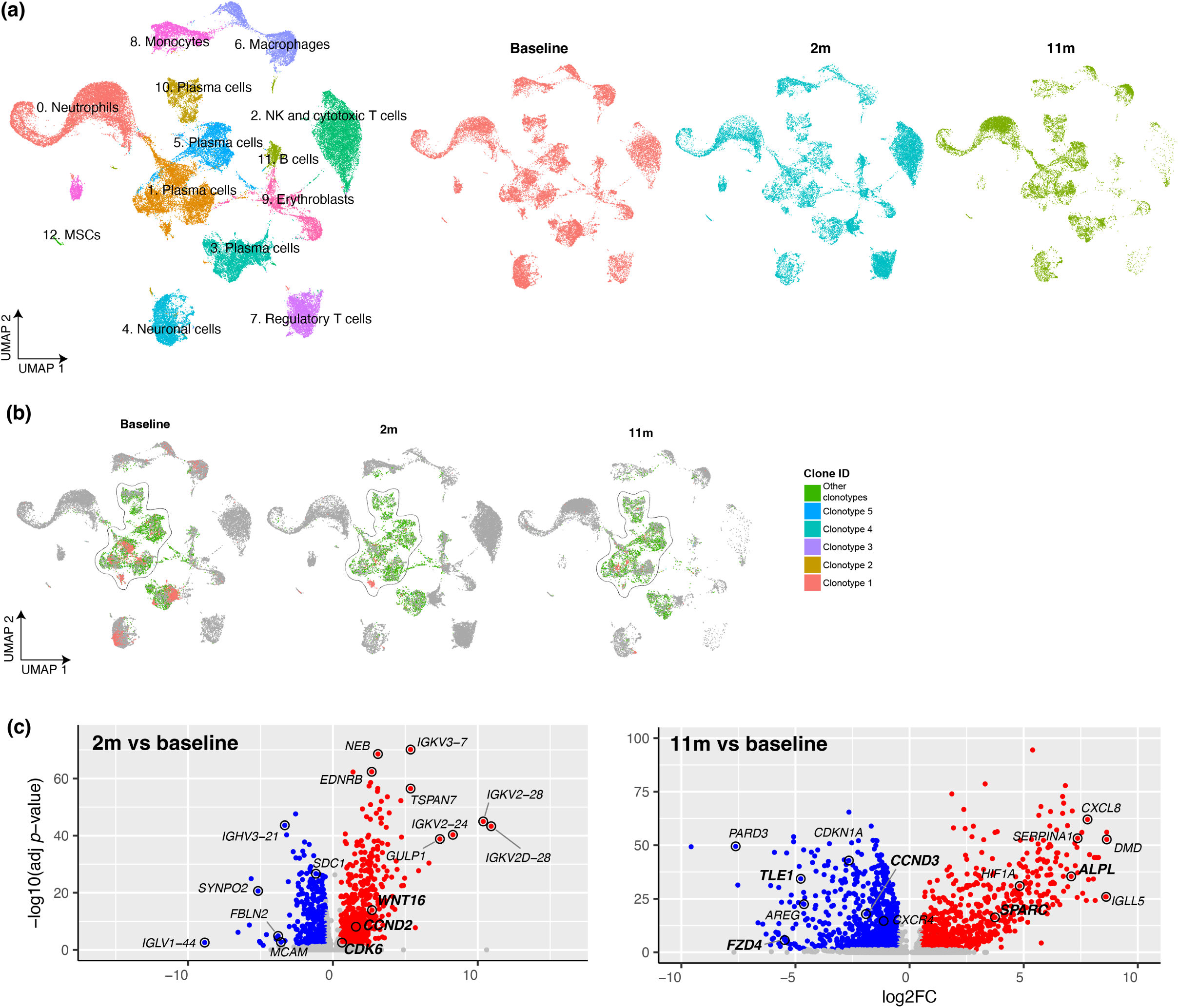
Effect of Romosozumab treatment on myeloma cells in aspirate **(a)** UMAP plot showing cell clusters identified in the aspirate across all treatment timepoints. **(b)** Mapping of distinct plasma cell clonotypes across treatment timepoints. **(c)** Volcano plots showing DEGs within the dominant plasma cell clone (Clonotype 1) relative to baseline following 2 months and 11 months of romosozumab treatment. Genes in bold text are involved in or direct targets of Wnt pathway.

**Supplementary Figure 4:**
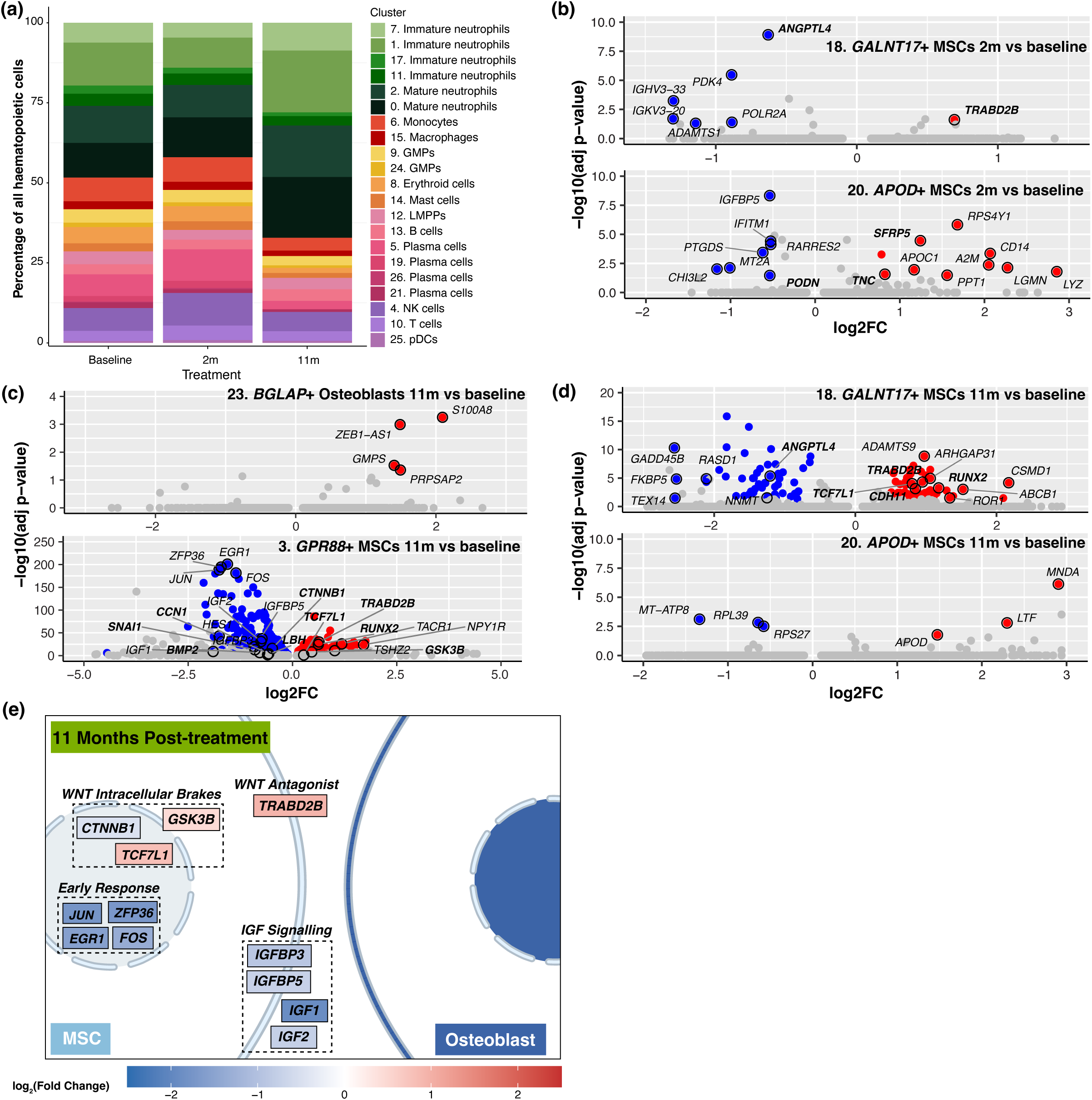
Effect of Romosozumab treatment on cell populations in bone trephines **(a)** Stacked bar plot illustrating the proportional distribution of hematopoietic cell types across treatment timepoints. **(b–d)** Volcano plots showing differentially expressed genes (DEGs) in MSC and osteoblast clusters relative to baseline: **(b)** Cluster 18 *GALNT17*^+^ MSCs (top) and Cluster 20 *APOD*^+^ MSCs (bottom) at 2 months; **(c)** Cluster 23 *BGLAP*^+^ osteoblasts (top) and Cluster 3 *GPR88*^+^ MSCs (bottom) at 11 months; and **(d)** Cluster 18 *GALNT17*^+^ MSCs (top) and Cluster 20 *APOD*^+^ MSCs (bottom) at 11 months. Genes in bold text are involved in or direct targets of Wnt pathway. **(e)** Schematic summarizing coordinated transcriptional changes within MSCs and osteoblasts following 11 months of romosozumab treatment.

## REFERENCES

1 Terpos, E. et al. Treatment of multiple myeloma-related bone disease: recommendations from the Bone Working Group of the International Myeloma Working Group. The Lancet Oncology 22, e119–e130 (2021). 10.1016/S1470-2045(20)30559-3

2 Kyle, R. A. et al. Review of 1027 Patients With Newly Diagnosed Multiple Myeloma. Mayo Clinic Proceedings 78, 21–33 (2003). 10.4065/78.1.21

3 Thorsteinsdottir, S. et al. Fractures and survival in multiple myeloma: results from a population-based study. Haematologica 105, 1067–1073 (2020). 10.3324/haematol.2019.230011

4 Morgan, G. J. et al. First-line treatment with zoledronic acid as compared with clodronic acid in multiple myeloma (MRC Myeloma IX): a randomised controlled trial. The Lancet 376, 1989–1999 (2010). 10.1016/S0140-6736(10)62051-X

5 Raje, N. et al. Evaluating results from the multiple myeloma patient subset treated with denosumab or zoledronic acid in a randomized phase 3 trial. Blood Cancer Journal 6, e378 (2016). 10.1038/bcj.2015.96

6 Raje, N. et al. Denosumab versus zoledronic acid in bone disease treatment of newly diagnosed multiple myeloma: an international, double-blind, double-dummy, randomised, controlled, phase 3 study. The Lancet Oncology 19, 370–381 (2018). 10.1016/S1470-2045(18)30072-X

7 Tian, E. et al. The Role of the Wnt-Signaling Antagonist DKK1 in the Development of Osteolytic Lesions in Multiple Myeloma. New England Journal of Medicine 349, 2483–2494 (2003). 10.1056/NEJMoa030847

8 Oshima, T. et al. Myeloma cells suppress bone formation by secreting a soluble Wnt inhibitor, sFRP-2. Blood 106, 3160–3165 (2005). 10.1182/blood-2004-12-4940

9 Giuliani, N. et al. Production of Wnt Inhibitors by Myeloma Cells: Potential Effects on Canonical Wnt Pathway in the Bone Microenvironment. Cancer Research 67, 7665–7674 (2007). 10.1158/0008-5472.CAN-06-4666

10 Giuliani, N. et al. Myeloma cells block RUNX2/CBFA1 activity in human bone marrow osteoblast progenitors and inhibit osteoblast formation and differentiation. Blood 106, 2472–2483 (2005). 10.1182/blood-2004-12-4986

11 Ehrlich, L. A. et al. IL-3 is a potential inhibitor of osteoblast differentiation in multiple myeloma. Blood 106, 1407–1414 (2005). 10.1182/blood-2005-03-1080

12 Adamik, J. et al. EZH2 or HDAC1 Inhibition Reverses Multiple Myeloma–Induced Epigenetic Suppression of Osteoblast Differentiation. Molecular Cancer Research 15, 405–417 (2017). 10.1158/1541-7786.MCR-16-0242-T

13 Garcia-Gomez, A. et al. Targeting aberrant DNA methylation in mesenchymal stromal cells as a treatment for myeloma bone disease. Nature Communications 12, 421 (2021). 10.1038/s41467-020-20715-x

14 Yaccoby, S. et al. Antibody-based inhibition of DKK1 suppresses tumor-induced bone resorption and multiple myeloma growth in vivo. Blood 109, 2106–2111 (2006). 10.1182/blood-2006-09-047712

15 Heath, D. J. et al. Inhibiting Dickkopf-1 (Dkk1) Removes Suppression of Bone Formation and Prevents the Development of Osteolytic Bone Disease in Multiple Myeloma. Journal of Bone and Mineral Research 24, 425–436 (2009). 10.1359/jbmr.081104

16 Fulciniti, M. et al. Anti-DKK1 mAb (BHQ880) as a potential therapeutic agent for multiple myeloma. Blood 114, 371–379 (2009). 10.1182/blood-2008-11-191577

17 Chantry, A. D. et al. Inhibiting activin-A signaling stimulates bone formation and prevents cancer-induced bone destruction in vivo. Journal of Bone and Mineral Research 25, 2633–2646 (2010). 10.1002/jbmr.142

18 Vallet, S. et al. Activin A promotes multiple myeloma-induced osteolysis and is a promising target for myeloma bone disease. Proceedings of the National Academy of Sciences 107, 5124–5129 (2010). 10.1073/pnas.0911929107

19 Raje, N. & Vallet, S. Sotatercept, a soluble activin receptor type 2A IgG-Fc fusion protein for the treatment of anemia and bone loss. Curr Opin Mol Ther 12, 586–597 (2010).

20 Terpos, E. et al. Elevated circulating sclerostin correlates with advanced disease features and abnormal bone remodeling in symptomatic myeloma: reduction post-bortezomib monotherapy. International Journal of Cancer 131, 1466–1471 (2012). 10.1002/ijc.27342

21 Gerov, V. et al. Dynamics of Bone Disease Biomarkers Dickkopf-1 and Sclerostin in Patients with Multiple Myeloma. Journal of Clinical Medicine 12, 4440 (2023).

22 McDonald, M. M. et al. Inhibiting the osteocyte-specific protein sclerostin increases bone mass and fracture resistance in multiple myeloma. Blood 129, 3452–3464 (2017). 10.1182/blood-2017-03-773341

23 Delgado-Calle, J. et al. Genetic deletion of Sost or pharmacological inhibition of sclerostin prevent multiple myeloma-induced bone disease without affecting tumor growth. Leukemia 31, 2686–2694 (2017). 10.1038/leu.2017.152

24 Sabol, H. M. et al. Healing of lytic lesions and restoration of bone health in multiple myeloma through sclerostin inhibition. Experimental Hematology & Oncology 14, 108 (2025). 10.1186/s40164-025-00699-4

25 McClung Michael, R., et al. Romosozumab in Postmenopausal Women with Low Bone Mineral Density. New England Journal of Medicine 370, 412–420 10.1056/NEJMoa1305224

26 Cosman, F. et al. Romosozumab Treatment in Postmenopausal Women with Osteoporosis. New England Journal of Medicine 375, 1532–1543 10.1056/NEJMoa1607948

27 Saag Kenneth, G., et al. Romosozumab or Alendronate for Fracture Prevention in Women with Osteoporosis. New England Journal of Medicine 377, 1417–1427 10.1056/NEJMoa1708322

28 Palla, B., Anderson, J., Miloro, M., Moles, S. & Callahan, N. Romosozumab-associated medication-related osteonecrosis of the jaw. Oral and Maxillofacial Surgery Cases 9, 100318 (2023). 10.1016/j.omsc.2023.100318

29 Cosman, F. et al. Romosozumab Treatment in Postmenopausal Women with Osteoporosis. New England Journal of Medicine 375, 1532–1543 (2016). 10.1056/NEJMoa1607948

30 Saag Kenneth, G., et al. Romosozumab or Alendronate for Fracture Prevention in Women with Osteoporosis. New England Journal of Medicine 377, 1417–1427 (2017). 10.1056/NEJMoa1708322

31 McClung Michael, R., et al. Romosozumab in Postmenopausal Women with Low Bone Mineral Density. New England Journal of Medicine 370, 412–420 (2014). 10.1056/NEJMoa1305224

32 Langdahl, B. L. et al. Romosozumab (sclerostin monoclonal antibody) versus teriparatide in postmenopausal women with osteoporosis transitioning from oral bisphosphonate therapy: a randomised, open-label, phase 3 trial. The Lancet 390, 1585–1594 (2017). 10.1016/S0140-6736(17)31613-6

33 Durie, B. G. M. et al. International uniform response criteria for multiple myeloma. Leukemia 20, 1467–1473 (2006). 10.1038/sj.leu.2404284

34 Humbert, L. et al. 3D-DXA: Assessing the Femoral Shape, the Trabecular Macrostructure and the Cortex in 3D from DXA images. IEEE Transactions on Medical Imaging 36, 27–39 (2017). 10.1109/TMI.2016.2593346

35 Lewiecki, E. M. et al. 3D-modeling from hip DXA shows improved bone structure with romosozumab followed by denosumab or alendronate. Journal of Bone and Mineral Research 39, 473–483 (2024). 10.1093/jbmr/zjae028

36 Winzenrieth, R. et al. Abaloparatide Effects on Cortical Volumetric BMD and Estimated Strength Indices of Hip Subregions by 3D-DXA in Women With Postmenopausal Osteoporosis. Journal of Clinical Densitometry 25, 392–400 (2022). 10.1016/j.jocd.2021.11.007

37 Mann, D. C. et al. Evaluating Skellytour for Automated Skeleton Segmentation from Whole-Body CT Images. Radiology: Artificial Intelligence 7, e240050 (2025). 10.1148/ryai.240050

38 Isensee, F., Jaeger, P. F., Kohl, S. A. A., Petersen, J. & Maier-Hein, K. H. nnU-Net: a self-configuring method for deep learning-based biomedical image segmentation. Nature Methods 18, 203–211 (2021). 10.1038/s41592-020-01008-z

39 Chai, R. C. et al. Multiscale analysis and functional validation of the cellular and genetic determinants of skeletal disease. Nature Genetics (In Press).

40 Lopes, M. D. et al. Romosozumab in postmenopausal women with smoldering multiple myeloma: a prospective 12-mo study. JBMR Plus 9, ziaf144 (2025). 10.1093/jbmrpl/ziaf144

41 Ominsky, M. S., Niu, Q. T., Li, C., Li, X. & Ke, H. Z. Tissue-level mechanisms responsible for the increase in bone formation and bone volume by sclerostin antibody. Journal of Bone and Mineral Research 29, 1424–1430 (2014). 10.1002/jbmr.2152

42 Eriksen, E. F. et al. Modeling-Based Bone Formation After 2 Months of Romosozumab Treatment: Results From the FRAME Clinical Trial. Journal of Bone and Mineral Research 37, 36–40 (2022). 10.1002/jbmr.4457

43 Eriksen, E. F. et al. Reconstruction of remodeling units reveals positive effects after 2 and 12 months of romosozumab treatment. Journal of Bone and Mineral Research 39, 729–736 (2024). 10.1093/jbmr/zjae055

44 Lewiecki, E. M. et al. One Year of Romosozumab Followed by Two Years of Denosumab Maintains Fracture Risk Reductions: Results of the FRAME Extension Study. Journal of Bone and Mineral Research 34, 419–428 (2019). 10.1002/jbmr.3622

45 Cosman, F., McMahon, D., Dempster, D. & Nieves, J. W. Standard Versus Cyclic Teriparatide and Denosumab Treatment for Osteoporosis: A Randomized Trial. Journal of Bone and Mineral Research 35, 219–225 (2020). 10.1002/jbmr.3850

46 Leder, B. Z. et al. Denosumab and teriparatide transitions in postmenopausal osteoporosis (the DATA-Switch study): extension of a randomised controlled trial. The Lancet 386, 1147–1155 (2015). 10.1016/S0140-6736(15)61120-5

47 Constantinescu, C. S., Farooqi, N., O’Brien, K. & Gran, B. Experimental autoimmune encephalomyelitis (EAE) as a model for multiple sclerosis (MS). British Journal of Pharmacology 164, 1079–1106 (2011). 10.1111/j.1476-5381.2011.01302.x

48 Hao, Y. et al. Dictionary learning for integrative, multimodal and scalable single-cell analysis. Nature Biotechnology 42, 293–304 (2024). 10.1038/s41587-023-01767-y

49 Korsunsky, I. et al. Fast, sensitive and accurate integration of single-cell data with Harmony. Nature Methods 16, 1289–1296 (2019). 10.1038/s41592-019-0619-0

50 Street, K. et al. Slingshot: cell lineage and pseudotime inference for single-cell transcriptomics. BMC Genomics 19, 477 (2018). 10.1186/s12864-018-4772-0

